# Women’s views and experiences of accessing vaccination in pregnancy during the COVID-19 pandemic: A multi-methods study in the United Kingdom

**DOI:** 10.1101/2021.09.14.21263505

**Authors:** Helen Skirrow, Sara Barnett, Sadie Bell, Sandra Mounier-Jack, Beate Kampmann, Beth Holder

## Abstract

**Background:** COVID-19 changed access to healthcare, including vaccinations, in the United Kingdom (UK). This study explored UK women’s experiences of accessing pertussis vaccination during pregnancy and infant vaccinations during COVID-19.

**Methods:** An online cross-sectional survey was completed, between 3^rd^ August-11 ^th^ October 2020, by 1404 women aged 16+ years who were pregnant at some point after the first UK lockdown from March 23^rd^, 2020. Ten follow-up semi-structured interviews were conducted.

**Results:** Most women surveyed were pregnant (65.7%) and a third postnatal (34.3%). Almost all women (95.6%) were aware that pertussis vaccination is recommended in pregnancy. Most pregnant (72.1%) and postnatal women (84.0%) had received pertussis vaccination; however, access issues were reported.

Over a third (39.6%) of women had a pregnancy vaccination appointment changed. COVID-19 made it physically difficult to access pregnancy vaccinations for one fifth (21.5%) of women and physically difficult to access infant vaccinations for almost half of women (45.8%). Nearly half of women (45.2%) reported feeling less safe attending pregnancy vaccinations and over three quarters (76.3%) less safe attending infant vaccinations due to COVID-19. The majority (94.2%) felt it was important to get their baby vaccinated during COVID-19.

Pregnant women from ethnic-minorities and lower-income households were less likely to have been vaccinated. Minority-ethnicity women were more likely to report access problems and feeling less safe attending vaccinations for both themselves and their babies.

Qualitative analysis found women experienced difficulties accessing antenatal care and relied on knowledge from previous pregnancies to access vaccines in pregnancy.

**Conclusion:** During the ongoing and future pandemics, healthcare services should prioritise equitable access to routine vaccinations, including tailoring services for ethnic-minority families who experience greater barriers to vaccination.

**Highlights:**

- Access to pregnancy vaccines in the United Kingdom was disrupted by the COVID-19 pandemic.
- UK women reported difficulties in physically accessing vaccine appointments and feeling less safe accessing vaccine appointments for themselves when pregnant and for their babies during COVID-19, with women from ethnic minorities in the UK were more likely to report difficulties.
- Vaccine services must ensure equitable access to vaccine appointments during the ongoing COVID-19 pandemic including tailoring services for lower income and ethnic minority families.

## Background

The COVID-19 pandemic has affected the delivery and uptake of routine vaccination programmes for pregnant women and children worldwide (1-6) increasing the risk of infectious disease outbreaks (7). In the United Kingdom (UK), despite routine vaccination programmes for pregnant women and children being prioritised (8) by the National Health Service (NHS), changes to primary-care (9) and maternity services (10) during the pandemic disrupted vaccination programme (7, 11). For example, decreased numbers of measles, mumps and rubella (MMR) vaccines were delivered after the start of the first national lockdown in March 2020 in England (12). Declines in childhood vaccination in England recovered over the summer of 2020; however, overall childhood vaccine uptake in England for 2020 was lower compared to previous years (13). In the UK, pregnant women are routinely advised to be vaccinated against pertussis (whooping cough) between 16 and 32 weeks of pregnancy, as well as seasonal influenza. Pertussis vaccine uptake for pregnant women in England eligible to be vaccinated after the first national lockdown in 2020 was 4.1% lower than the same period in 2019; the lowest it has been since 2016 (14).

Traditionally, vaccinations are delivered in primary care via general practices (GPs) in the UK; however some hospital-based antenatal clinics also offer vaccines to pregnant women (15). During the first national lockdown the public were advised to *‘stay at home’* and avoid attending healthcare settings unless necessary. A reduction in primary care use was observed (16) and routine vaccines services adapted in primary care (17). The shift to more remote consultations and reductions in face-to-face appointments (9, 10), may have reduced women’s access to vaccines.

Work by co-authors Bell et al. found that UK parents felt confused about whether routine childhood vaccine services were still running normally during the first national lockdown (18). Pregnant women were classified as at increased risk from COVID-19 and advised by the UK government to adhere closely to social distancing guidance (19). These messages may have therefore affected pregnant women’s willingness to attend healthcare facilities for vaccination, as seen for childhood vaccination (3, 18). Two qualitative studies by Anderson et al have reported that pregnant women felt confused by the risk COVID-19 represented to them and that some women also factored the risk of COVID-19 when considering attending routine pregnancy vaccine appointments (20, 21). There has yet to be a detailed report on the impact of the COVID-19 pandemic on women’s access to routine pregnancy vaccines.

This multi-methods study investigates the impact of the COVID-19 pandemic on women’s awareness, and acceptance of pertussis vaccine in pregnancy and access to pertussis vaccine and childhood vaccines in the UK.

## Methods

A multi-methods approach was taken – using quantitative and qualitative components – with the aim of gaining insight into women’s awareness of pertussis vaccination and access to pregnancy and childhood vaccines during COVID-19. The study comprised of an online survey and semi-structured interviews, to both quantify different experiences of accessing routine vaccines in pregnancy and then explore in greater depth barriers and facilitators to accessing routine vaccines during the COVID-19 pandemic. Due to the timing of the onset of the pandemic and the first national lockdown in the UK, this study focuses on pertussis vaccination in pregnancy.

### Survey design

The survey was designed based on previous surveys on pregnancy vaccination (22) and through consultation and piloting among midwives, pregnancy vaccine researchers, paediatricians and public health professionals. The survey gathered optional demographic data including ethnicity, age, number of children, country of residence, region of residence (England participants only), parity, income, pregnancy status, gestation at survey completion for those who were pregnant, and date of delivery for those who had already had their babies.

### Survey Content

The survey included questions about pertussis vaccine awareness and uptake during pregnancy. Questions also asked about access to antenatal care and the pertussis vaccination during the pandemic, including where they would have preferred to receive their vaccine and what sources of information on pregnancy vaccines they would prefer. Questions also asked about whether the COVID-19 pandemic had restricted their physical access to vaccines or made them feel less safe getting themselves and their baby vaccinated. Women were also asked whether they felt it was important to have their baby vaccinated during the pandemic. See supplementary file A for full survey.

### Ethical approval

This study was approved by Imperial College Research Ethics Committee (ICREC) (Ref: 20IC6188).

### Survey Recruitment

The survey was advertised and promoted using Facebook with a landing page and paid advertising using Facebook’s ad manager which crossposts to Instagram. The three adverts had a combined reach of 46,146, with 1,573 post engagements and 1,394 link clicks. Related organisations on Facebook were also contacted individually by study researchers, including pregnancy yoga and birth preparation classes, breastfeeding support groups and toddler groups. The survey was shared and distributed via the research team’s personal twitter accounts, including linking to other researchers and organisations with maternal and vaccine uptake interests. Finally, the survey was also promoted via some Maternity Voices Partnerships (23) who were e-mailed and invited to share the survey, and via a post on the website Mumsnet.

Eligible participants were required to have been pregnant at some point between the start of the first UK lockdown (from 23rd March 2020) and the time of survey completion, to be resident in the UK, and to be aged 16 years or over. The survey was live from 3^rd^ August until 11^th^ October 2020. The online survey was prefaced by an information page explaining the study, and how the data was to be used (Supplementary material A). Participants were informed that by taking part in the survey they agreed for their responses to be used for research purposes. Participants were required to confirm (by tick-box) at the start of the survey that they met the eligibility criteria and that they consented to participate in the survey.

### Survey Analysis

Descriptive statistics of survey respondents were reported, and statistical analysis undertaken using Stata (version-16). Binary answers, including awareness and uptake of pertussis vaccination, were compared by age group, household income, ethnicity, region and country of the United Kingdom using Pearson’s chi-squared statistic.

For responses to questions on Likert scales (e.g., where responses were scored on a scale of *‘Strongly agree, somewhat agree, neither agree nor disagree, somewhat disagree, strongly disagree and not/applicable’)*, stepwise logistic regression was undertaken to determine factors associated with the responses. To facilitate analysis with the number of responses obtained, the <20y group was combined with the next age bracket to form < 25y age bracket, the ten income groups were combined pairwise to form five groups, and ethnicity was dichotomised into ‘White’ (i.e., White British, White Irish and White Other participants) and ‘Ethnic minorities’ (i.e., Black, Asian, Chinese, Mixed ethnicities or Other ethnicities). A p value of less than 0.05 was considered statistically significant.

### Semi-structured interviews

At the end of the survey, participants were invited to take part in a follow-up interview by leaving their contact details. A selection of participants who had left their contact details were contacted by SBa. Participants were purposively selected to prioritise women: 1) from ethnic-minority backgrounds, due to lower representation among survey respondents; 2) who were pregnant when surveyed, due to their proximity to their pregnancy experience compared to those that had already had their babies ; 3) who had not completed the open text responses. Informed written and verbal consent was obtained from participants. The participant information sheet and consent form were provided by e-mail (Supplementary material B). Interviews lasted approximately 30 minutes and were conducted over the telephone and/or using Microsoft Teams and were recorded with permission of the participant. Interviews were conducted by SBa and HS using a topic guide. The topic guide was developed based on the questionnaire (Supplementary material C). The interviews took place between the 7^th^ and 16th of December 2020.

### Qualitative analysis

Qualitative interviews and free-text survey responses were analysed thematically by SBa using the stages outlined by Braun and Clarke: data familiarisation, coding and theme identification and refinement (24). To enhance the rigour of the analysis, coding approaches and subsequent theme generation and refinement was discussed between HS, SBa, SBe and BH.

## Results

### Demographics

In total, our recruitment strategy led to 1526 survey click throughs. 122 responses were excluded because they were pilot responses or incomplete responses, leaving 1,404 responses. Available demographic details are summarised in Supplementary Figure 1. Most women were 30-34 years (n=548, 39.0%; Supplementary Figure 1A), and White British (n=1072, 76.4%; Supplementary Figure 1B). Most women were pregnant at the time of taking the survey (n=922, 65.7%; Supplementary Figure 1C). Median household annual income was £ 45,999-54,999 (Supplementary Figure 1D) and most women worked full-time (n=848, 60.4%; Supplementary Figure 1E). Most women either had no other children (n=517, 36.8%) or one child (n=524, 37.3%; Supplementary Figure 1F). The majority were living in England (n=1289, 91.8%; Supplementary Figure 1G), and the highest proportion were from London and the South-West (Supplementary Figure 1H). Women that had given birth by the time of survey completion had mostly delivered between April and August 2020 (Supplementary Figure 1I). Those who were pregnant at the time of survey completion ranged from 5 to 41 weeks’ gestation, with 34 weeks being the most frequent gestation (n=50; Supplementary Figure 1J). The characteristics of the women interviewed are shown in Supplementary Table 1.

### Findings

Findings from the quantitative survey analysis, and the qualitative analysis of the free-text survey responses and the interviews are presented together. To support findings the overarching themes from the qualitative analysis, with supporting quotes, are provided in Table 1.

**Table 1:**
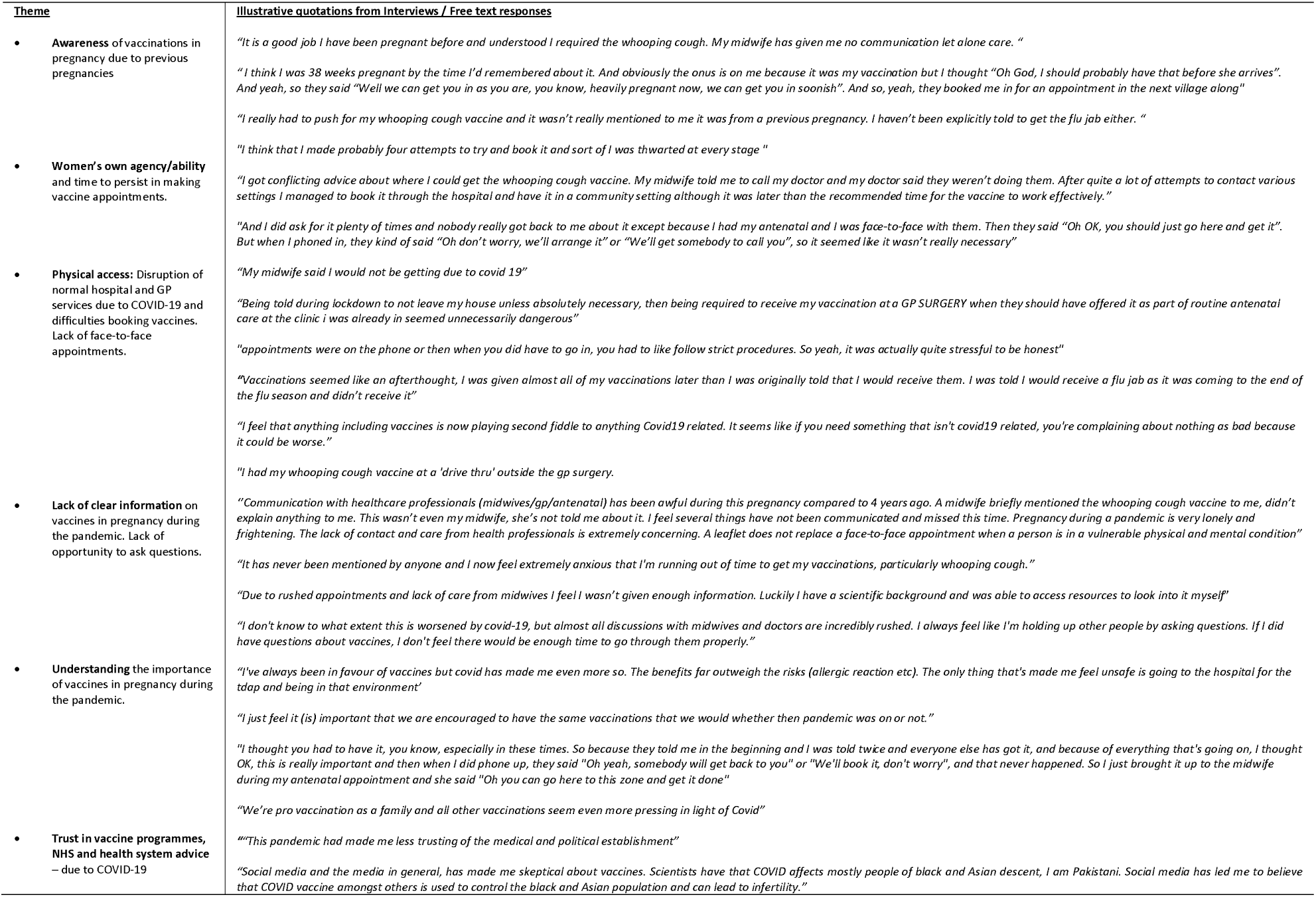
Interview and free text responses illustrating themes

### Awareness of pertussis vaccine recommendation in pregnancy

The majority of women (95.6%, n=1338) – including those pregnant and those postnatal - were aware that pertussis vaccination is recommended in pregnancy (Figure 1A). Women with an annual household income below £25,000 were more likely to be unaware (p=0.02) of pertussis vaccination being recommended in pregnancy (Figure 1B). Awareness of pertussis vaccine in pregnancy did not otherwise vary by age, ethnicity or geographically (Table 2). Qualitative analysis found that women’s awareness of the pertussis vaccination often came from their own prior personal pregnancy experiences (Table 1).

**Figure 1:**
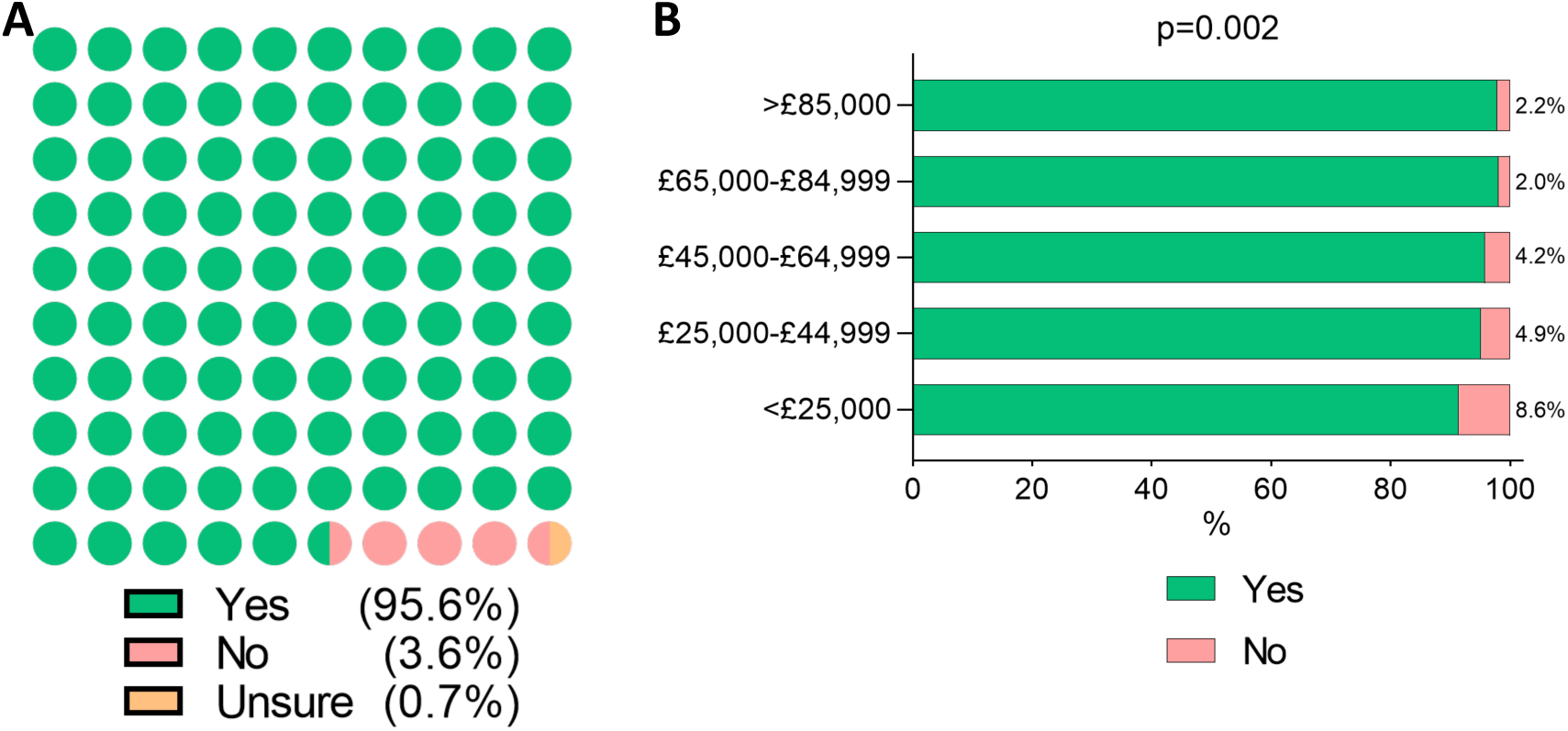
Awareness that pertussis vaccination is recommended in pregnancy during the COVID-19 pandemic. Participants were asked the question: ‘Whooping cough vaccine (also known as pertussis or Tdap) is recommended to all pregnant people in the UK. Were you aware of this?’. Possible answers were: ‘Yes’, ‘No’ or ‘Unsure’. A) Awareness among all survey respondents at the time of survey completion (both women who were currently pregnant and new mothers). B) Awareness of respondents separated by income bracket. Chi-Square Test of association between annual household income and pertussis vaccine awareness; p = <0.002.

**Table 2:** Awareness of pertussis vaccine being recommended in pregnancy in the UK

| Whooping cough vaccine (also known as pertussis or Tdap) is recommended to all pregnant people in the UK. Were you aware of this? |  |  |  |
| --- | --- | --- | --- |
|  | YES | NO | Chi square test of independence |
|  | n (%) | n (%) |  |
| All women | 1,338 (95.6) | 61 (4.4) | - |
| **Ethnicity** |  |  |  |
| White | 1,019 (95.5) | 48 (4.5) | $\chi^2 = 0.86$ |
| Minority ethnicity | 101 (93.5) | 7 (6.5) | $p = 0.353$ |
| **Income** |  |  |  |
| Under £24,999 | 137 (89.5) | 16 (10.5) | $\chi^2 = 20.13$<br>$p = <0.001$ |
| £25,000-£44,999 | 250 (95.1) | 13 (4.9) |  |
| £45,000-£64,999 | 339 (95.8) | 15 (4.2) |  |
| £65,000-£84,999 | 239 (97.9) | 5 (2.1) |  |
| Over £85,000 | 261 (97.8) | 6 (2.2) |  |
| **Age** |  |  |  |
| Under 25y | 84 (92.1) | 7 (7.7) | $\chi^2 = 6.22$<br>$p = 0.183$ |
| 25 – 29y | 334 (96.5) | 12 (3.5) |  |
| 30 – 34 y | 524 (95.8) | 22 (4.0) |  |
| 35 – 39 y | 292 (96.1) | 12 (3.9) |  |
| Over 39y | 65 (91.6) | 6 (8.4) |  |
| **Country** |  |  |  |
| Scotland | 51 (98.1) | 1 (1.9) | $\chi^2 = 2.09$<br>$p = 0.552$ |
| Wales | 35 (94.6) | 2 (5.4) |  |
| Northern Ireland | 26 (100) | 0 (0) |  |
| England | 1,226 (95.5) | 58 (4.5) |  |
| **Region** |  |  |  |
| Greater London | 180 (95.2) | 9 (4.8) | $\chi^2 = 9.13$<br>$p = 0.332$ |
| East Midlands | 74 (94.9) | 4 (5.1) |  |
| West Midlands | 107 (97.3) | 3 (2.7) |  |
| North East | 58 (98.3) | 1 (1.7) |  |
| South East | 224 (96.6) | 8 (3.4) |  |
| South West | 95 (91.4) | 9 (8.6) |  |
| Yorkshire and the Humber | 209 (94.1) | 13 (5.9) |  |
| East of England | 109 (97.3) | 3 (2.7) |  |

Women were asked the question: 'Whooping cough vaccine (also known as pertussis or Tdap) is recommended to all pregnant people in the UK. Were you aware of this?'. Possible answers were: 'Yes', 'No' or 'Unsure'. No and unsure combined into NO.
**Ethnicity Groups: White:** White-British, White-Irish, White-Other; **Minority ethnicity:** Black-British African, Black-British Caribbean and Black-Other, Asian Indian, Asian-Pakistani, Asian-Bangladeshi, Asian-Other and Chinese, Mixed White-Black Caribbean, Mixed White-Black African, Mixed White-Asian, Mixed White/Other and Other ethnicity.

### Pertussis vaccine information for pregnant women during COVID-19

Women surveyed showed a preference for receiving vaccine information from midwives and GPs (Figure 2A) and preferred face-to-face information above other types of information such as leaflets (Figure 2B). Qualitative analysis found women commonly reported a lack of opportunity to get information about vaccines in pregnancy, associated with wider issues around a lack of general information provided in pregnancy, compared to their experiences of previous pregnancies prior to lockdown: *“Communication with healthcare professionals (midwives/GP/antenatal) has been awful during this pregnancy compared to 4 years ago……The lack of contact and care from health professionals is extremely concerning. A leaflet does not replace a face-to-face appointment when a person is in a vulnerable physical and mental condition”*.

**Figure 2:**
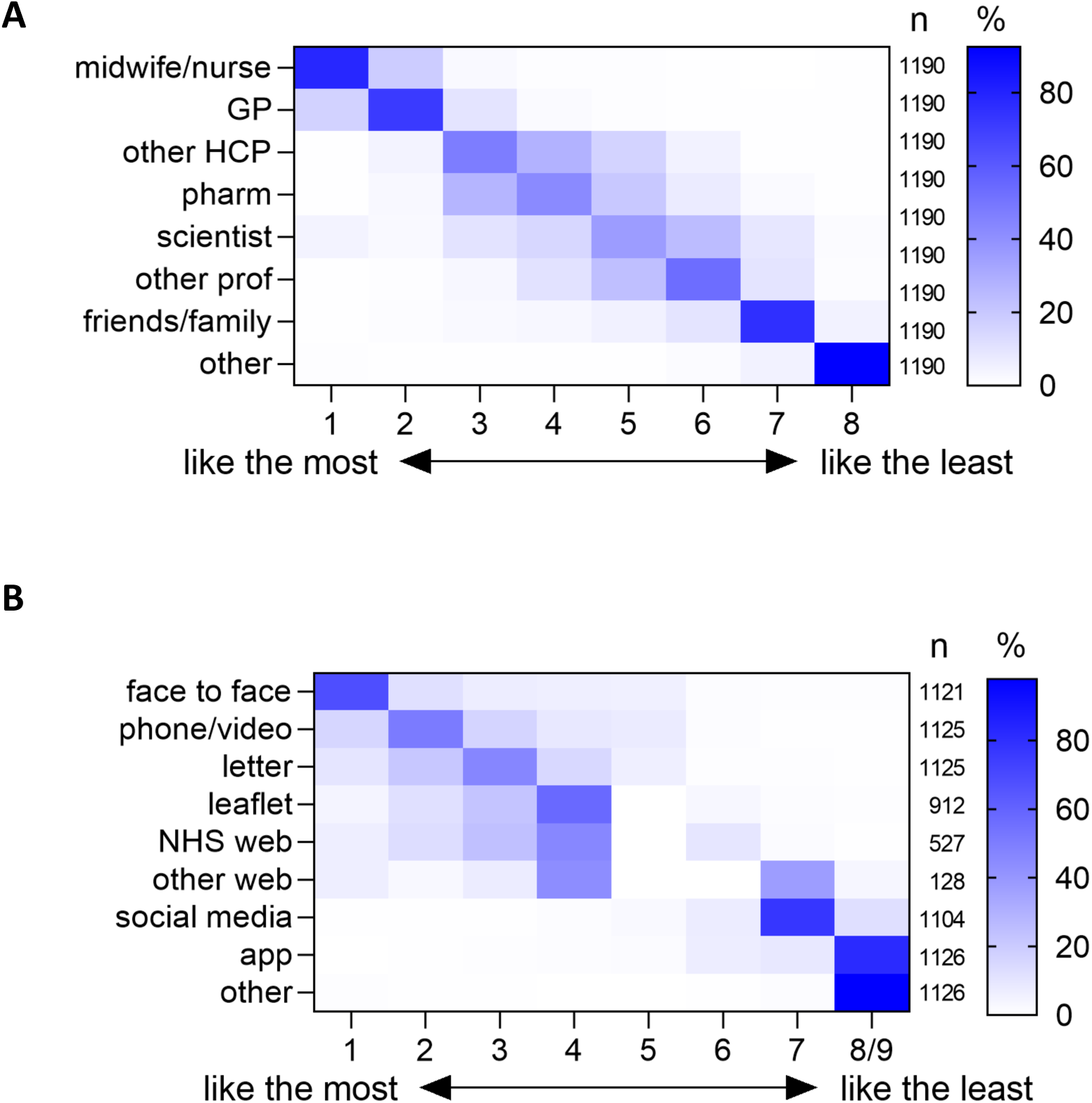
Preferred information sources regarding vaccination in pregnancy during the COVID-19 pandemic. A) The question posed was ‘Who would you like to get information from about vaccinations in pregnancy during the coronavirus pandemic?’ and respondents asked to rank the listed options from the one they liked the most (1) to the one they liked the least (8). B) The question posed was ‘How would you like to get information from about vaccinations in pregnancy during the coronavirus pandemic?’ and respondents asked to rank the options from the one they liked the most (1) to the one they liked the least (9). GP; general practitioner, other HCP; other healthcare professional, other prof; other professional; web; website

### Uptake of pertussis vaccine in pregnancy during COVID-19

Uptake of pertussis vaccine during pregnancy among women who were currently pregnant was 72.1% (n=624) and among new mothers was 89.1% (n=408) (Supplementary Table 2). Among women who were currently pregnant and unvaccinated against pertussis (n=241) the majority were in the first or second trimester (82.6%, n=199) (Supplementary Table 2) and most were intending to be vaccinated (84.6%, n=204) (Supplementary Table 3).

Among women who were pregnant when surveyed but unaware that pertussis vaccination was recommended (n=46), when asked *‘Now you know that this vaccine is recommended in pregnancy, do you think you will get vaccinated?*’ most (76%) answered that they would or were leaning towards being vaccinated (Supplementary Table 4).

Of women vaccinated against pertussis in a previous pregnancy (n=188) most were also vaccinated in this pregnancy (n=181, Figure 3). Only 6.9% of women changed to not being vaccinated in pregnancy. 30% of women who were unvaccinated in their most recent previous pregnancy were vaccinated in this pregnancy during COVID-19 (Figure 3).

**Figure 3:**
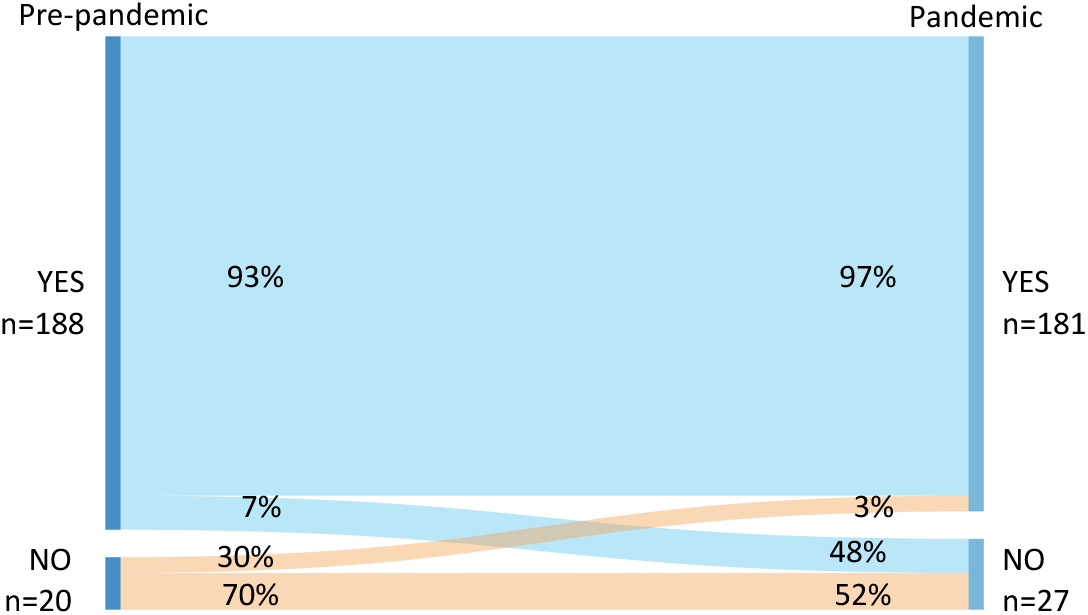
Pertussis vaccination uptake in pregnancies during the COVID-19 pandemic compared to pre-pandemic pregnancies. Sankey plot showing pertussis vaccination uptake in women who had been previously pregnant in the last eight years (left; pre-pandemic), compared to their pregnancy during the pandemic (right). n= 208. Diagram created using SankeyMATIC.

Qualitative analysis found that some women felt that vaccination in pregnancy during the pandemic was even more important than ever as a motivator for getting vaccinated: *“I have always believed in vaccines but covid has made me even more so*.*”* However, women also considered the risk of COVID-19 when attending vaccine appointments such as: *“The only thing that’s made me feel unsafe is going to the hospital for the tdap* (pertussis vaccine) *and being in that environment”*. Some women also described feeling that their trust in either vaccines or the wider medical establishment was influenced by the pandemic for example one women said, *“This pandemic had made me less trusting of the medical and political establishment”*. One woman who self-reported as Pakistani described how social media had led her to believe that *“COVID vaccine amongst others is used to control the black and Asian population and can lead to infertility”*.

### Predictors of pertussis vaccination uptake in pregnancy during COVID-19

Pregnant women in the first and second trimester had lower vaccine uptake compared to women in their third trimester (Supplementary Table 2) however, most planned to be vaccinated (Supplementary Table 3). Therefore, to look at predictors of vaccination among pregnant women, we looked just at those who were in their third trimester. Women in their third trimester who were from an ethnic-minority (excluding White ethnic-minorities) were more likely to be unvaccinated compared to White ethnicity women (p=0.041; Table 3). Those aged under 25 years were also more likely to be unvaccinated compared to those aged 30-34 years (p=<0.0001; Table 3). Income did not predict vaccine uptake in these women (p=0.884; Table 3). Among new mothers, women from ethnic minorities (p=0.001) were more likely to be unvaccinated compared to women of White ethnicity (Table 3). In this group, living in households with an income below £ 25,000 was also associated with higher likelihood to be unvaccinated (p=0.02; Table 3). In these women, age did not predict vaccine uptake (p>0.05; data not shown).

**Table 3:**
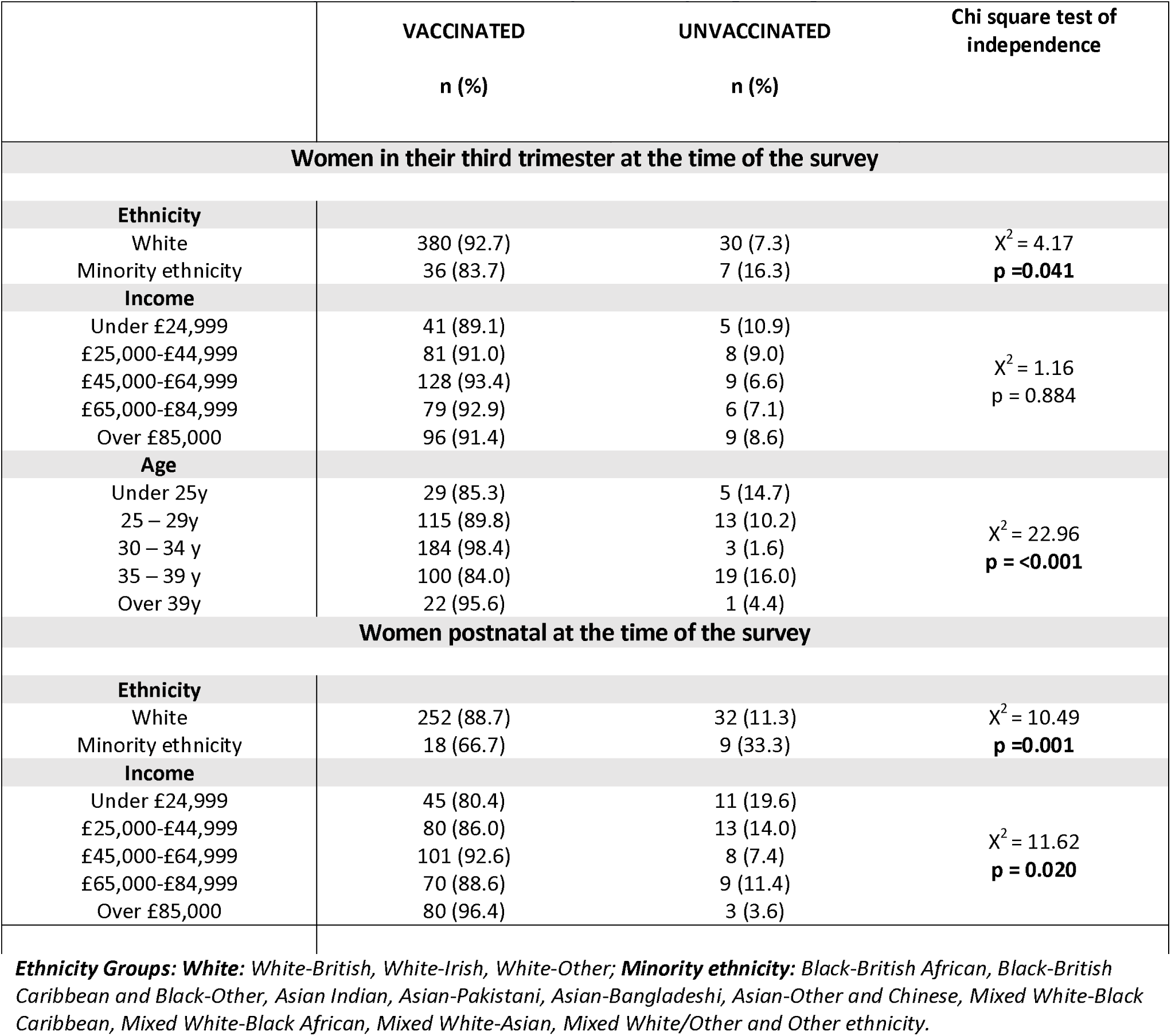
Predictors of pertussis vaccine uptake in pregnancy

Multivariate logistic regression that considered ethnicity, age and income, found that for women in their third trimester, most age groups were more likely to be unvaccinated compared to the most common 30–34-year age group, and age and ethnicity were no longer significant predictors. In women who had already delivered, being unvaccinated was still associated with being from an ethnic-minority (p=<0.001) and a lower-income household (<£ 25,000, p=0.012; Table 4).

**Table 4:** Multivariate analysis of predictors of pertussis vaccine uptake

| Variable | Pregnant at time of survey |  |  | Delivered at the time of survey |  |  |
| --- | --- | --- | --- | --- | --- | --- |
|  | OR | 95% CI | P value | OR | 95% CI | P value |
| **Ethnicity** |  |  |  |  |  |  |
| White# | - | - | - | - | - | - |
| Minority ethnicity | 2.26 | 0.82, 6.25 | 0.116 | 8.33 | 2.53, 27.49 | **<0.001** |
| **Age** |  |  |  |  |  |  |
| Under 25y | 6.23 | 1.13, 34.23 | **0.035** | 1.89 | 0.55, 6.50 | 0.706 |
| 25 – 29y | 5.26 | 1.34, 20.57 | **0.017** | 0.27 | 0.07, 1.03 | 0.636 |
| 30 – 34 y # | - | - | - | - | - | - |
| 35 – 39 y | 8.79 | 2.45, 31.49 | **0.001** | 0.74 | 0.25, 2.19 | 0.075 |
| Over 39y | 2.21 | 0.21, 22.97 | 0.508 | 1.64 | 0.36, 7.48 | 0.484 |
| **Income** |  |  |  |  |  |  |
| Under £24,999 | 0.77 | 0.19, 3.09 | 0.712 | 7.71 | 1.56, 38.16 | **0.012** |
| £25,000-£44,999 | 0.95 | 0.31, 2.90 | 0.935 | 3.61 | 0.75, 17.34 | 0.109 |
| £45,000-£64,999 | 0.52 | 0.17, 1.58 | 0.249 | 2.65 | 0.51, 13.58 | 0.242 |
| £65,000-£84,999 | 0.77 | 0.23, 2.54 | 0.671 | 4.01 | 0.84, 18.98 | 0.080 |
| Over £85,000# | - | - | - | - | - | - |

**OR:** ordinal odds ratio. An OR above 1 indicates a higher likelihood of women being unvaccinated in pregnancy.
**95% CI:** 95% confidence interval. # indicates the comparator group in the analysis. **Ethnicity Groups: White:** White-British, White-Irish, White-Other; **Minority ethnicity:** Black-British African, Black-British Caribbean and Black-Other, Asian Indian, Asian-Pakistani, Asian-Bangladeshi, Asian-Other and Chinese, Mixed White-Black Caribbean, Mixed White-Black African, Mixed White-Asian, Mixed White/Other and Other ethnicity.

Among unvaccinated pregnant women (n=87), 25% (n=22) answered that ‘they wanted to be vaccinated but didn’t because of the COVID-19 pandemic’ (Supplementary Table 5). Among unvaccinated women who had already delivered (n=47), only 2% (n=1) answered that they ‘didn’t want to be vaccinated’, 43% (n=20) ‘didn’t know about the vaccine’ and 21% (n=10), answered that ‘they wanted to be vaccinated but didn’t because of the COVID-19 pandemic’ (Supplementary Table 5).

### Access to pregnancy vaccine appointments during COVID-19

Among the >1,000 women surveyed the majority (62%) had experienced a GP appointment for their pregnancy being changed, cancelled or postponed due to COVID-19, with appointments most commonly being changed to phone or online appointments (53.3%) (Table 5). There were more reports of appointments being postponed or cancelled by the GP (10.8% and 16.2% respectively), compared to then being postponed or cancelled by patients (2.1% and 1.1% respectively). Over a third of women (39.6%) surveyed reported that their GP appointment for a vaccine was changed, postponed or cancelled during the pandemic. Only 7.5% of women reported that a vaccine hospital appointment had been changed. Nearly half (42.9%) reported that a hospital appointment for another reason had been changed, postponed or cancelled. Most of these (33.2%) were changed to phone or online appointments (Table 5).

**Table 5:** Appointment changes during COVID-19 pandemic

| n=1404 | GP apt<br>for pregnancy | GP apt<br>for vaccine | GP apt for other | Hospital apt<br>for vaccine | Hospital apt<br>for other |
| --- | --- | --- | --- | --- | --- |
| **Postponed by patient** | 2.1% | 1.4% | 0.8% | 0.5% | 1.5% |
| **Postponed by GP/hospital** | 10.8% | 2.8% | 2.6% | 2.7% | 7.3% |
| **Cancelled by patient** | 1.1% | 1.1% | 0.6% | 0.6% | 0.5% |
| **Cancelled by GP/hospital** | 16.2% | 2.6% | 3.9% | 2.3% | 9.1% |
| **Changed to phone/online** | 53.3% | 1.6% | 33.8% | 1.7% | 33.2% |
| **Blank (no disruptions/NA)** | 533 | 848 | 1275 | 1298 | 801 |
| **Total reports disruptions** | 68.8% | 13.6% | 31.5% | 7.7% | 47.5% |
| **% patients reporting disruption** | 62.0% | 39.6% | 9.2% | 7.5% | 42.9% |

**Women were asked 'Have you had any appointments changed due to the COVID-19 pandemic'** for the following appointment types. 'An appointment at your GP surgery about your pregnancy', 'An appointment at your GP surgery to receive a pregnancy vaccine', 'An appointment at your GP for another reason', 'An appointment at a hospital to receive a pregnancy vaccine' and 'An appointment at the hospital for another reason'. **Apt = appointment.**

Qualitative analysis found that the onus was on the women to organise and push to have their vaccines in pregnancy and first-time parents who didn’t have the prior knowledge about vaccines in pregnancy found it particularly difficult. For example, one woman said: *“It is a good job I have been pregnant before and understood I required the whooping cough. My midwife has given me no communication”*.

### Physical access and safety attending pregnancy vaccine appointments

Among all women 21.5% (n=256) strongly agreed or somewhat agreed that COVID-19 had restricted their physical access to pregnancy vaccines (Figure 4A). Nearly half of women (45.2%, n=564) strongly agreed (12.8%) or somewhat agreed (32.4%) that the pandemic had made them feel less safe attending their pregnancy vaccine appointments (Figure 4B).

**Figure 4.**
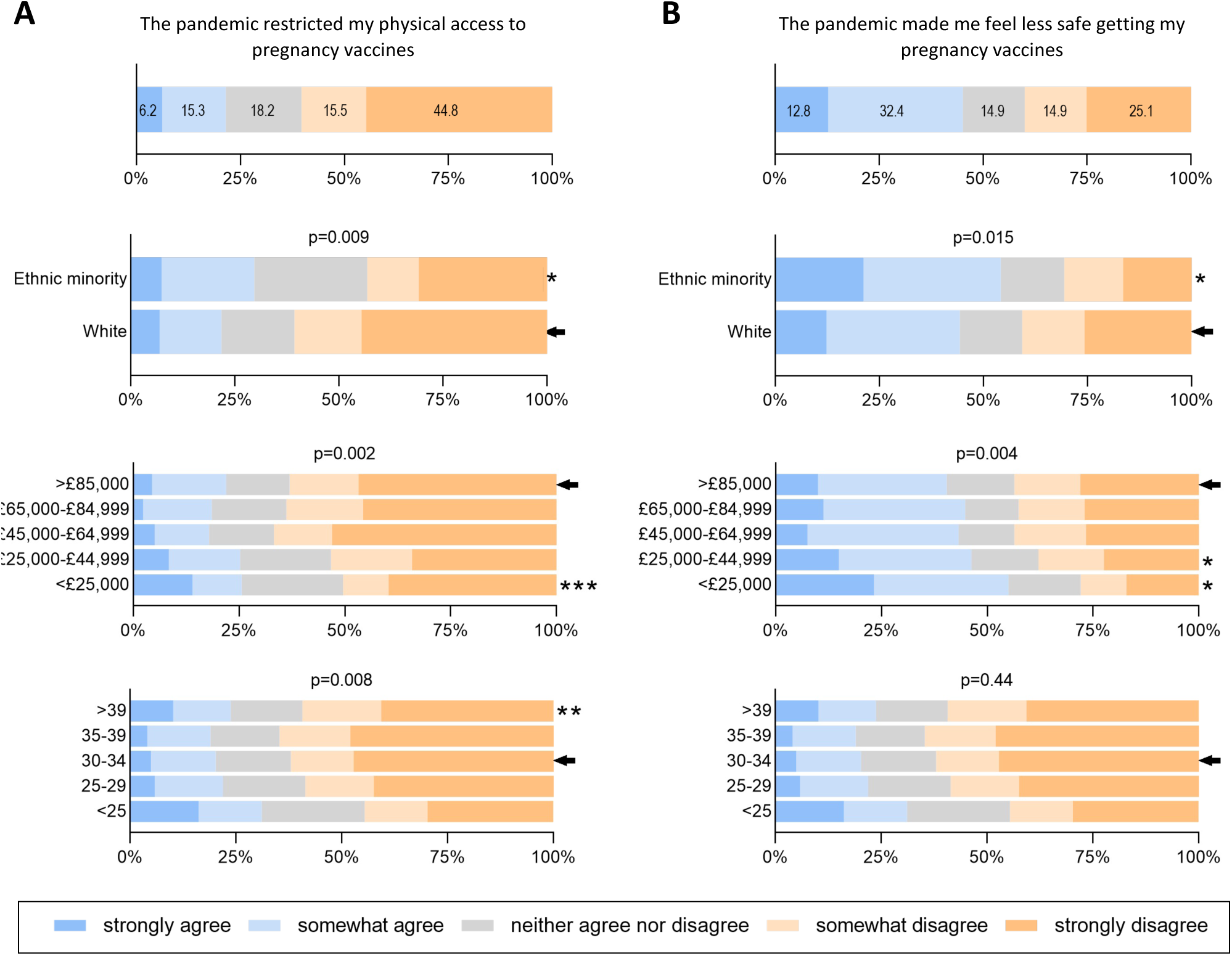
Perceptions of physical restriction and safety when attending for vaccination during pregnancy. A) Women were asked to what extent they agreed with the following statement : “The COVID pandemic has restricted my physical access to vaccines during pregnancy”, and to record this on a Likert scale (see key). The top plot shows all respondents (n=1,191). The lower plots shows responses by ethnicity (with respondents separated into ethnic minority (n=81) and white ethnicity (n=929) groups), by income and by age. **B)** Women were asked to what extent they agreed with the following statement: “The COVID pandemic has restricted my physical access to vaccines during pregnancy” and to record this on a Likert scale (see key). The top plot shows all respondents (n=1,249). The lower plots shows responses by ethnicity (with respondents separated into ethnic minority (n=81) and white ethnicity (n=929) groups), by income and by age. Differences were analysed using ordered logistic regression. The comparator group is indicated by the arrow, and significant differences indicated with asterisks. * p<0.05, ** p<0.01, *** p<0.001.

We next looked at the predictors of the responses to the questions about physical restriction and feeling safe getting pregnancy vaccines. Women who were from ethnic minorities other than White were more likely to report that the pandemic had restricted their physical access (Figure 4A, p=0.009) and that they felt less safe attending for pregnancy vaccines (Figure 4B, p=0.015), compared to women of White ethnicities. Women from lower-income households (<£ 25,000) were more likely to have felt their access to pregnancy vaccine appointments was restricted or felt less safe attending vaccine appointments due to COVID-19 compared to the highest income households (Figures 4A & 4B). Women aged under 25 years were also more likely to have felt their access to pregnancy vaccine appointments was restricted (Figure 4A);however, they did not report a higher feeling less safe attending vaccine appointments due to COVID-19 (Figure 4B).

Qualitative analysis also found that women reported confusion and difficulties in booking vaccine appointments in pregnancy and a lack of available information about pregnancy vaccines during COVID-19. This meant some women experienced being vaccinated later in pregnancy than recommended. For example, *“I got conflicting advice about where I could get the whooping cough vaccine……After quite a lot of attempts to contact various settings I managed to book……and have it in a community setting although it was later than the recommended time for the vaccine to work effectively*.*”* Confusion was often related to wider disruption to all antenatal care appointments and not specific to pregnancy vaccine appointments. Women described the stress and anxiety caused by the move from face-to-face to remote appointments in antenatal care and COVID-19 protocols: *“appointments were on the phone or then when you did have to go in, you had to like follow strict procedures. So yeah, it was actually quite stressful to be honest”*.

### Physical access, safety and importance of vaccinating their baby during COVID-19

The majority of women (94.2%, n=1,106) felt that it was important to get their baby vaccinated during the COVID-19 pandemic (Figure 5A, 85.9% strongly and 8.3% somewhat agreeing). The majority (76.3%, n=893) of women agreed strongly (44.2%) or somewhat agreed (32.0%) that they felt safe to go and get their baby vaccinated during the pandemic (Figure 5A). Nearly half (45.8%) of women strongly agreed or somewhat agreed that the COVID-19 pandemic would make it physically difficult to get their baby vaccinated (Figure 5A).

**Figure 5:**
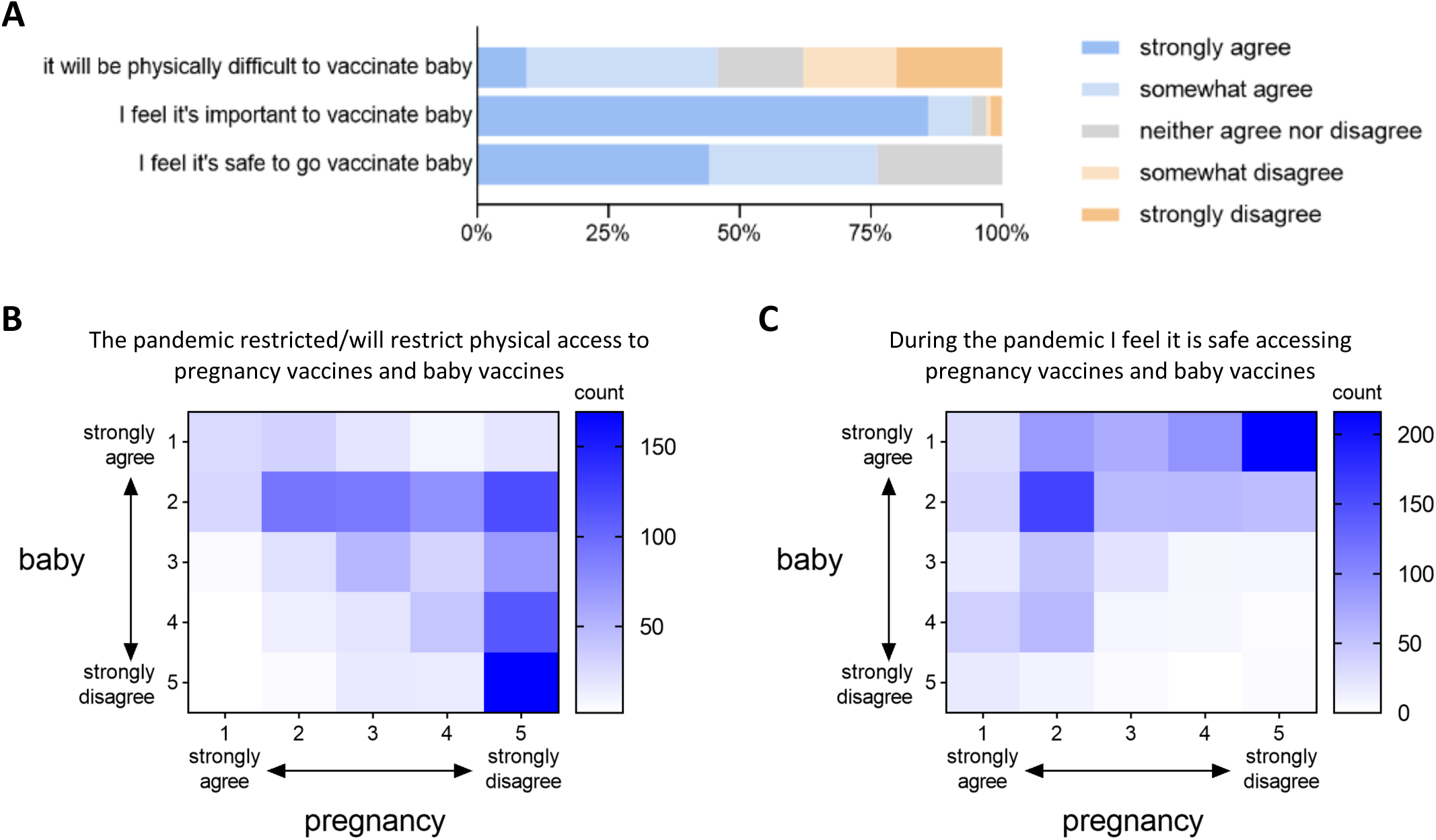
Women’s perceptions of physical restriction, importance, and safety regarding attending vaccination appointments for their babies, compared to during pregnancy. A: Women were asked to what extent they agreed with the following statements, recording their responses on a Likert scale: ‘The COVID-19 pandemic will make it physically difficult to get my baby vaccinated’, ‘During the COVID-19 pandemic I feel it is important to get my baby vaccinated’ and ‘During the COVID-19 pandemic I feel it is safe about to go to get my baby vaccinated”. B) Heat map showing the relationship between women’s perceptions of COVID-19 making it physically difficult to get vaccinated during pregnancy (columns) compared to it making it physically difficult to get their baby vaccinated (rows). C) Heat map showing the relationship between feeling safe accessing pregnancy vaccines during the COVID-19 pandemic (columns) and feeling safe getting baby vaccinated during the COVID-19 pandemic (rows). 1: *strongly disagree. 2: somewhat disagree, 3: neither agree nor disagree, 4: somewhat agree, 5: strongly agree*.

We next performed multivariate analyses to investigate predictors of responses to infant vaccination, considering ethnicity, age and income. With age and income considered, belonging to an ethnic-minority was predictive of women feeling that they would find it physically difficult to access vaccines for their babies (p=0.001), feeling less safe accessing vaccines for their baby (p=0.001) and reporting lower importance to vaccinating their baby during the pandemic (p=<0.001) (Table 6). Women aged under 25 years were more likely to feel less safe getting their baby vaccinated (p=0.001) (Table 6). Women from lower-income households (<£ 45,000) were more likely to report lower importance for vaccinating their baby during the pandemic compared to the highest income group. Income was not a predictor in multi-variate analysis for feeling less safe or finding getting their baby vaccinated during COVID-19 was physically harder (Table 6).

**Table 6:** Multivariate analysis of predictors of attitudes to accessing to baby vaccinations

| Variable | The COVID-19 pandemic will make it **physically difficult** to get my baby vaccinated |  |  | During the COVID-19 pandemic I feel it is **important** to get my baby vaccinated |  |  | During the COVID-19 pandemic I feel it is **safe** to go to get my baby vaccinated |  |  |
| --- | --- | --- | --- | --- | --- | --- | --- | --- | --- |
|  | OR | 95% CI | P value | OR | 95% CI | P value | OR | 95% CI | P value |
| **Ethnicity** |  |  |  |  |  |  |  |  |  |
| White# | - | - | - | - | - | - | - | - | - |
| Minority ethnicity | 0.46 | 0.29, 0.74 | **0.001** | 3.41 | 1.95, 5.95 | **<0.001** | 2.14 | 1.36, 3.39 | **0.001** |
| **Age** |  |  |  |  |  |  |  |  |  |
| Under 25y | 0.67 | 0.39, 1.16 | 0.155 | 1.25 | 0.59, 2.62 | 0.550 | 1.69 | 1.00, 2.88 | **0.049** |
| 25 – 29y | 1.18 | 0.87, 1.61 | 0.281 | 0.99 | 0.58, 1.66 | 0.968 | 0.91 | 0.66, 1.25 | 0.56 |
| 30 – 34 y # | - | - | - | - | - | - | - | - | - |
| 35 – 39 y | 0.98 | 0.72, 1.34 | 0.902 | 0.88 | 0.49, 1.54 | 0.648 | 0.88 | 0.64, 1.22 | 0.46 |
| Over 39y | 1.14 | 0.67, 1.94 | 0.638 | 1.30 | 0.58, 2.95 | 0.525 | 1.46 | 0.83, 2.55 | 0.19 |
| **Income** |  |  |  |  |  |  |  |  |  |
| Under £24,999 | 1.00 | 0.63, 1.59 | 0.982 | 3.97 | 1.99, 7.96 | **<0.001** | 1.57 | 0.97, 2.52 | 0.064 |
| £25,000-£44,999 | 0.82 | 0.56, 1.21 | 0.327 | 1.96 | 1.02, 3.79 | **0.044** | 1.19 | 0.80, 1.79 | 0.373 |
| £45,000-£64,999 | 1.14 | 0.80, 1.60 | 0.473 | 0.79 | 0.39, 1.62 | 0.535 | 0.99 | 0.69, 1.42 | 0.963 |
| £65,000-£84,999 | 1.11 | 0.77, 1.59 | 0.556 | 1.49 | 0.77, 2.88 | 0.234 | 1.2 | 0.84, 1.79 | 0.283 |
| Over £85,000# | - | - | - | - | - | - | - | - | - |

Women were asked how much they agreed or disagreed with the following statements:
**OR:** ordinal odds ratio. An OR above 1 indicates a higher likelihood of women giving responses moving from *Strongly agree, somewhat agree, neither agree nor disagree, somewhat disagree, strongly disagree and not/applicable* so are more likely to have disagreed with the statement. **95% CI:** 95% confidence interval. # indicates the comparator group in the analysis. **Ethnicity Groups: White:** White-British, White-Irish, White-Other; **Minority ethnicity:** Black-British African, Black-British Caribbean and Black-Other, Asian Indian, Asian-Pakistani, Asian-Bangladeshi, Asian-Other and Chinese, Mixed White-Black Caribbean, Mixed White-Black African, Mixed White-Asian, Mixed White/Other and Other ethnicity.

Women who had answered that the COVID-19 pandemic had made their access to vaccines in pregnancy physically difficult were also likely to report that the COVID-19 pandemic meant they felt it would be physically difficult to get their baby vaccinated (Figure 5B). Women’s responses to feeling safe getting themselves vaccinated in pregnancy or getting their babies vaccinated were less aligned (Figure 5C).

### Delivery of pertussis vaccines during and prior to the COVID-19 pandemic

Of those vaccinated in pregnancy, most women had been vaccinated at their GP (n=637, 61.8%), followed by a hospital antenatal care setting (n=316, 30.7%) (Supplementary Table 6). Among unvaccinated pregnant women, the majority were similarly planning to attend either their GP (n=114, 55.9%) or a hospital antenatal care setting (n=59, 28.9%, supplementary Table 7).

Vaccination location varied by country and English region (Supplementary Figures 2A and 2B). More Scottish women were vaccinated in a hospital antenatal setting (42.9%, n=18), followed by English women (31.6%, n=298), whereas no Welsh nor Northern Irish women were vaccinated in hospital (Supplementary Figure 2A, p=<0.001). In Wales and Northern Ireland, vaccination by GPs was far higher; 92.1% and 100% in Wales and NI respectively. Location of vaccination also varied across regions of England (Supplementary Figure 2B) with 65.5% (n=38) of women in the East Midlands vaccinated at antenatal hospital settings compared to only 2.3% (n=1) of women in the North-East region of the UK, (p=<0.001). Vaccination in antenatal community settings was highest in the North-East and North-West England. Women’s preference for where they wanted to be vaccinated during pregnancy in the COVID-19 pandemic showed higher preference for GPs and community antenatal settings (Supplementary Figure 2C).

Most women who were vaccinated at a GP surgery in their most recent previous pregnancy were also vaccinated at a GP surgery during their COVID-19 pregnancy (Supplementary Figure 2D). The main change in location was that 46% (n=53) of women who were vaccinated at a hospital antenatal appointment during COVID-19 had been vaccinated at a GP surgery in their previous pregnancy (Supplementary Figure 2D).

### Travel to healthcare settings during COVID-19

Given government advice on travel and public transport use during the pandemic, women were asked how they usually travelled to various health care providers, before and during the pandemic. Travel by public transport to GPs, pharmacies and antenatal appointments all decreased during the pandemic (Supplementary Figure 3A); 40.1% women travelled using public transport to antenatal appointments before the pandemic, which dropped to only 6.2% during the pandemic (Supplementary Figure 3B). Public transport to attend antenatal appointments varied regionally, pre and post COVID-19 with women from London reporting highest pre-pandemic public transport use (40.1%) (supplementary Figure 1C) and also the greatest change (84.5% drop: supplementary Figures 3D & 3E).

## Discussion

Despite finding high awareness and uptake among the over 1000 women surveyed, we found that women’s access to pertussis vaccination during the pandemic was disrupted. Qualitative findings suggest that women’s previous knowledge and pregnancy experiences before COVID-19 highly influenced their vaccination experiences. Women described having to be proactive in getting vaccine appointments and confusion about how to access antenatal care. Nearly two thirds of women surveyed reported that their antenatal appointments had been changed due to COVID-19, most commonly to remote consultations.

The majority (95.6%) of women were aware that the pertussis vaccine is recommended in pregnancy. We found higher pertussis vaccine uptake (84%) among women surveyed who had already delivered their baby compared to national pertussis vaccine uptake estimates during the same time period (14). Among currently pregnant women surveyed uptake was lower (72.1%); however, the majority of pregnant women who were unvaccinated were in the first and second trimester and were planning to get vaccinated, so this lower uptake is likely due to their gestation when completing the survey.

The women surveyed and interviewed felt that their antenatal care had been impacted negatively by the pandemic including perceiving a lack of information about how to get vaccinated in pregnancy. The lack of face-to-face appointments was mentioned by different women as a key barrier to accessing information and advice from healthcare professionals. Appointments were described as rushed and women found being pregnant during COVID-19 stressful.

Our survey findings suggest that vaccination experiences varied in the UK by region, ethnicity, income, age and was also influenced by their previous pregnancies. Women from ethnic minorities were less likely to have been vaccinated in pregnancy and were also more likely to report feeling less safe attending vaccine appointments and that their access to vaccine appointments had been physically restricted due to the pandemic for them and their babies. Women from lower-income households were less likely to be aware of pertussis vaccine recommendation and remain unvaccinated by the time they delivered their baby. Younger women below the age of 25 were less likely to have been vaccinated in pregnancy, which may reflect them being pregnant for the first time as our qualitative findings highlight the importance of women’s prior knowledge from previous pregnancies. Women in lower-income groups were also more likely to think getting their baby vaccinated during COVID-19 was less important.

Our findings are consistent with studies prior to the pandemic with women living in poorer areas or belonging to an ethnic-minority being less likely to be vaccinated in pregnancy (15, 25, 26). This is mirrored in uptake of childhood vaccines in the UK, with lower-income and ethnic-minority children less likely to receive their routine vaccinations (25, 27, 28). Consistent with the findings by Bell et al., we also observed that women from ethnic minorities were more likely to report physical limitations and feeling unsafe when accessing vaccines for their children during the pandemic (18). Different factors affect ethnic minorities living the UK having lower vaccine uptake including access barriers and vaccine confidence (27, 28). However, during COVID-19 the increased risk to ethnic-minority populations has been well publicised (29) and ethnic-minority pregnant women are also more likely to admitted to hospital with COVID-19 (30). Ethnic-minority women may have therefore been more nervous about attending healthcare settings for pregnancy vaccines.

We found that more women were vaccinated at hospital antenatal settings during COVID-19 compared to previous pregnancies which supports previous work that antenatal hospital vaccine clinics play a key role in delivering pregnancy vaccines (15) and only 7.5% of women reported a hospital vaccine appointment being changed. Further evaluation of the impact of antenatally delivered vaccines on uptake, including during COVID-19 is required (14, 31). We also found that where women received pregnancy vaccines varied geographically across the UK nations and regionally. This could reflect variation in pre-existing maternal vaccine service provision prior to the pandemic, changes to services in response to the pandemic or transport variation. Mcquaid et al found that the impact of COVID-19 on routine childhood vaccine uptake varied between UK nations with England reporting a decline and Scotland an increase in childhood vaccine uptake in 2020 (11).

We found that public transport use to attend healthcare appointments decreased during COVID-19 and private car use increased, which was most marked in London, which had the highest pre-pandemic use of public transport. The impact of reduced public transport on healthcare access could be greater for people living on lower-incomes and belonging to ethnic minorities who are more likely to have fewer transport options available (32). This could explain why women from ethnic minorities were more likely to report that their physical access to vaccine appointments had been negatively impacted by COVID-19 given the government advice to avoid public transport and also the reduced public transport services available during lockdown.

Our findings add to the literature that routine care of pregnant women (10) and children (33), including routine vaccines has been impacted by COVID-19 (3, 12, 13, 18). Most women reported appointments being changed, cancelled and/or postponed, including for pregnancy vaccines and some also experienced being vaccinated later than recommended. In a study of 31 qualitative interviews with pregnant women, Anderson et al. did not find disruption to pregnancy vaccine appointments was a common theme, possibly due to the earlier timing of their interviews, in April and May 2020 (20). Consistent with our qualitative findings however, Anderson et al did report that some women debated the benefits of attending for vaccines in pregnancy versus their risk to COVID-19 exposure (20). They also found that pregnant women at the start of the pandemic were confused about social-distancing guidance (21). In a study looking at the impact of the pandemic on maternity services in general, Sanders et al reported that women felt anxious and confused by the changes to UK maternity services, with only 12.9% feeling virtual antenatal appointments met their needs and 37.6% unsure about accessing their GP (34). Similarly a national study of 477 families reported that women giving birth during the pandemic in England experienced poor communication resulting in them feeling anxious (35).

Our findings suggest difficulties in accessing vaccination appointments have been a more important barrier for pregnant women during COVID-19 than anti-vaccination sentiments. This is consistent with a pre-pandemic survey on childhood vaccination (28) and work by Edelstein et al which suggested that access is a larger barrier to vaccination than anti-vaccination sentiment (36). Healthcare providers in the UK must ensure that access to vaccine appointments is prioritised during COVID-19 for pregnant women and children (7). Our findings also suggest that COVID-19 made access more difficult for lower-income and ethnic-minority mothers, and therefore may have widened pre-existing inequalities in vaccine uptake (15, 27, 28, 37). Improving vaccine uptake among ethnic-minority and low-income women is paramount now given the inequalities experienced by UK low-income areas and ethnic minorities during COVID-19 (38).

Recommendation by a trusted healthcare provider has been found to be a key intervention to improve maternal and childhood vaccine uptake (15, 22, 39-41). Our findings support this, as women preferred vaccine information from healthcare professionals. Every contact with a pregnant woman should be seen as an opportunity to promote and remind about vaccines for both themselves and their children (28), particularly during the pandemic, and information must be accessible to all women (42).

Pregnant women are at greater risk of poor outcomes from COVID-19, and COVID-19 vaccines are recommended for UK pregnant women (43, 44). Our previously reported findings suggest that women from lower-income households and belonging to ethnic-minority groups are less likely to accept COVID-19 vaccines when pregnant, and therefore reflect existing pregnancy vaccine uptake inequalities (45). It is thus essential to focus on promoting COVID-19 vaccination in pregnancy alongside routine pregnancy vaccines, particularly among ethnic-minority and low-income women.

### Strengths and Limitations

The main strength of this study was the use of multiple methods – the qualitative analysis of the survey and interviews enabled factors behind the quantitative findings to be explored in detail. Our response rate of over 1000 UK women who had been pregnant during the first peak of the COVID-19 pandemic was excellent.

The survey included women from across the UK, enabling us to identify differences geographically however the majority of women were from England. Regionally, London and the South-East were overrepresented, but the survey included women with a range of ages and income levels and at different pregnancy gestations and both vaccinated and unvaccinated women. Ethnicity was dichotomised into White ethnicities and all other ethnicities for analysis, as we were underpowered to detect differences between individual ethnicity groups. The timing of the survey is a strength as it took place at a time of uncertainty for pregnant women and the healthcare system. Despite vaccines being prioritised for pregnant women, uptake has decreased since the start of the pandemic (14) and our findings therefore remain timely and relevant. However, repeating the survey now would be beneficial and a larger study may be able to detect additional regional and demographic vaccine service and uptake variation during COVID-19.

## Conclusion

Access to routine vaccinations for UK pregnant women and their babies was disrupted by COVID-19 and changes to antenatal care created confusion. Pre-existing pregnancy vaccine uptake inequalities may have been exacerbated by the pandemic with pregnant women living in lower-income households and belonging to ethnic minorities being less likely to be vaccinated and experiencing greater barriers to accessing vaccines. Our findings are relevant for the delivery and uptake of COVID-19 vaccines in pregnancy.

Vaccination in pregnancy and childhood must be prioritised during the ongoing pandemic. Healthcare services should provide pregnant women with clear, accessible information on the importance of vaccinations and how they can safely access vaccine appointments when pregnant and for their babies. Vaccine services must be equitable to optimise uptake among women from ethnic minorities and lower-income households.

## Data Availability

All data supporting this research publication is included in the manuscript and supplementary material.

## Acknowledgements

We would like to thank all survey and interview participants for their time to provide their views and opinions. Thank you to the Bay Wide, Royal Surrey, Sandwell and West Birmingham, Leicester City, Bradford, Bolton, Central Cheshire, Chester, Bath and North-East Somerset, Swindon and Wiltshire, Royal Free and Kingston Maternity Voices Partnerships who agreed to share the survey via their networks. Mr Toby Clements and Dr Thomas F Rice helped with creation and dissemination of the online survey. Thank you to Myrsini Kaforou for the travel heat maps.

## Author Contributions

HS contributed to research question formulation, funding acquisition, research design and carrying out the study, data curation, data analysis, writing original article draft and reviewing and editing.

SBa contributed to research question formulation, funding acquisition, design and carrying out the study, data curation, data analysis, writing original article draft and reviewing and editing.

SBe contributed to research question formulation, funding acquisition, research design and carrying out the study, data analysis and reviewing and editing final article.

LR contributed to research design and carrying out the study, data analysis and reviewing and editing final article.

SMJ contributed to research question formulation, funding acquisition, research design and reviewing and editing final article.

BK contributed to research question formulation, funding acquisition, research design and reviewing and editing final article.

BH contributed to research question formulation, funding acquisition, research design and carrying out the study, data curation, data analysis, writing original article draft and reviewing and editing.

BH, LR, HS and SBa have verified the underlying data.

