## Supplementary A for "Women’s views and experiences of accessing vaccination in pregnancy during the COVID-19 pandemic: A multi-methods study in the United Kingdom"

Start of Block: Background

Q62

**Your views on vaccinations in pregnancy during the coronavirus (COVID-19) pandemic** (V1.0 22/07/2020)

INVITATION
You are invited to take part in an anonymous survey looking at views on vaccines during pregnancy and experiences of accessing these vaccines during the SARS-COV-2 coronavirus (COVID-19) pandemic. We are asking people who have been pregnant at any point during the UK lockdown (since 23rd March 2020) to take part in our survey to help us identify ways of improving how pregnancy vaccinations are communicated and delivered during pandemics.

You are eligible to participate in this survey if you: - live in the UK
- are aged 16 years or older
- are currently pregnant, or were pregnant at any point during the UK lockdown (since 23rd March 2020)

**INVESTIGATORS**
Imperial College London, The Institute of Reproductive and Developmental Biology:
Beth Holder, *Lecturer in Maternal and Fetal Health*
Helen Skirrow, *Clinical Research Fellow in Public Health*
Sara Barnett, *Research Midwife*
London School of Hygiene and Tropical Medicine, The Vaccine Centre/Department of Public Health and Policy:
Beate Kampmann, *Professor in Paediatric Infection and Immunity*
Sandra Mounier-Jack, *Associate Professor in Health Policy*
Sadie Bell, *Research Fellow in Public Health Evaluation*

**ABOUT THIS SURVEY**
This research is funded by the Imperial College COVID Research Fund. It has received approval from the Imperial College London Research Ethics Committee (Reference 20IC6188). The survey will take around 5-10 minutes to complete.

**Do I have to take part?**
No. Your participation is completely voluntary, and you do not need to answer all the questions if you do not wish to. You may withdraw from the survey at any point during the survey by closing the browser window.

**Will my answers remain confidential?**

The survey is anonymous, and we will not ask for any information in the survey that could identify who you are. Giving your consent means that we may use your answers (but not your name) in reports, presentations and papers about the research. We will not store your IP address.

The data will be stored securely in password-protected files at Imperial College London and London School of Hygiene and Tropical Medicine. Anonymous data from this survey may be made available in a public data repository for use by other researchers. The findings from this study will be published in scientific papers, that will be free to access. Updates for participants will be posted on The Vaccine Centre website and the lead investigator's website (www.theholderlab.com/info).

At the end of the survey you will be given the opportunity to further voice your experiences and views regarding vaccines in pregnancy during the coronavirus (COVID-19) pandemic through a telephone or Skype interview with one of the study investigators. If you decide you would like to assist further in a telephone or video-based interview with our research midwife, you can provide your contact details for us to get in touch with you. If you provide your contact details, your completed survey will no longer be anonymous.

**For more information about this research please feel free to contact us at any time and we will be happy to answer your questions. You can reach us at:**

Sara Barnett (research midwife):.
Beth Holder (lead investigator)

These contact details are included again at the end of the survey.

**Just here out of curiosity?**

If you would like a copy of the questions we ask in this survey, please email Beth Holder on. Alternatively, if you enter the survey out of curiosity and are not eligible, please just put a note somewhere to indicate that you are just having a look at it. Thank you!

---

*

Q60 I confirm that:

- ☐ I live in the UK (1)
- ☐ I am 16 years old or above (2)
- ☐ I am currently pregnant, or have been pregnant at some point during the UK lockdown (since 23rd March 2020) (3)
- ☐ I consent to participate in this survey (4)

---

Q1 How old are you?

- ○ under 20 years (1)
- ○ 20-24 years old (2)
- ○ 25-29 years old (3)
- ○ 30-34 years old (4)
- ○ 35-39 years old (5)
- ○ 40 years and over (6)

---

Q2 Where do you live?

- ○ Scotland (1)
- ○ Wales (2)
- ○ Northern Ireland (3)
- ○ England (4)

---

*Display This Question:*

*If Where do you live? = England*

Q62 In which region of England do you live?

- ○ Greater London (1)
- ○ East of England (11)
- ○ East Midlands (2)
- ○ West Midlands (3)
- ○ North East (4)
- ○ North West (5)
- ○ South East (7)
- ○ South West (8)
- ○ Yorkshire and the Humber (9)
- ○ Prefer not to say (10)

---

Q3 Which of the following best describes your ethnicity?

▼ White British (1) ... Do not wish to say (17)

---

Q4 Which of the following best describes your employment status over the last year?

▼ Full-time employment (1) ... Do not wish to say (9)

---

Q63 What is your total household income in GBP (£) from all sources, before tax

▼ Under £15,000 (1) ... Prefer not to answer (11)

---

Q5 How many children are you the parent or guardian of?

- ○ 0 (1)
- ○ 1 (2)
- ○ 2 (3)
- ○ 3 (4)
- ○ 4 or more (5)

End of Block: Background

Start of Block: Access to healthcare

Q7 Before the lockdown began (23rd March 2020), how would you usually get to the following face-to-face appointments:

| | Walk (1) | Public transport (2) | Private car (3) | Taxi (4) | Not applicable- no appointment needed (6) |
|---|---|---|---|---|---|
| Your GP (1) | ○ | ○ | ○ | ○ | ○ |
| A pharmacy (2) | ○ | ○ | ○ | ○ | ○ |
| Your antenatal appointments (3) | ○ | ○ | ○ | ○ | ○ |

---

Q49 After the lockdown began (23rd March 2020), how have you usually got to the following:

| | Walk (1) | Public transport (2) | Private car (3) | Taxi (4) | Not applicable- no appointment needed (6) |
|---|---|---|---|---|---|
| Your GP (1) | ○ | ○ | ○ | ○ | ○ |
| A pharmacy (2) | ○ | ○ | ○ | ○ | ○ |
| Your antenatal appointments (3) | ○ | ○ | ○ | ○ | ○ |

Q24 Which describes you:

○ I am currently pregnant (1)

○ I was pregnant at some point during the lockdown (23rd March 2020 onwards) (2)

**End of Block: Access to healthcare**

**Start of Block: Currently pregnant block (branch displayed if currently pregnant selected)**

Q25 How many weeks pregnant are you?

________________________________________________________________

Q47 Whooping cough vaccine (also known as pertussis or Tdap) is recommended to all pregnant people in the UK. Were you aware of this?

- ◯ Yes (1)
- ◯ No (2)
- ◯ Unsure (4)

---

*Display This Question:*

*If Whooping cough vaccine (also known as pertussis or Tdap) is recommended to all pregnant people in... = No*

*Or Whooping cough vaccine (also known as pertussis or Tdap) is recommended to all pregnant people in... = Unsure*

Q58 Now you know that this vaccine is recommended in pregnancy, do you think you will get vaccinated?

- ◯ Yes (1)
- ◯ Unsure but leaning towards yes (2)
- ◯ Unsure but leaning towards no (3)
- ◯ No (4)

---

*Display This Question:*

*If Whooping cough vaccine (also known as pertussis or Tdap) is recommended to all pregnant people in... = Yes*

Q46 Have you been vaccinated against whooping cough (also known as pertussis or Tdap) during your pregnancy?

- ◯ Yes (1)
- ◯ No (2)

---

*Display This Question:*

*If Have you been vaccinated against whooping cough (also known as pertussis or Tdap) during your pre... = No*

Q47 Do you plan to be vaccinated against whooping cough (also known as pertussis or Tdap) during your pregnancy?

- ◯ Yes (1)
- ◯ Unsure but leaning towards yes (2)
- ◯ Unsure but leaning towards no (3)
- ◯ No (4)

---

*Display This Question:*

*If Do you plan to be vaccinated against whooping cough (also known as pertussis or Tdap) during your... = Unsure but leaning towards yes*

*Or Do you plan to be vaccinated against whooping cough (also known as pertussis or Tdap) during your... = Unsure but leaning towards no*

*Or Do you plan to be vaccinated against whooping cough (also known as pertussis or Tdap) during your... = No*

*Or Now you know that this vaccine is recommended in pregnancy, do you think you will get vaccinated? = Unsure but leaning towards yes*

*Or Now you know that this vaccine is recommended in pregnancy, do you think you will get vaccinated? = Unsure but leaning towards no*

*Or Now you know that this vaccine is recommended in pregnancy, do you think you will get vaccinated? = No*

Q58 Which of the following best describes why you are unsure, or not planning, to be vaccinated?

- ◯ Because of the coronavirus pandemic (give details below if you wish) (1) ________________________________________________
- ◯ For another reason (give details below if you wish) (2) ________________________________________________

---

*Display This Question:*

*If Do you plan to be vaccinated against whooping cough (also known as pertussis or Tdap) during your... = No*

Q59 Which of the following best describes why you do not plan to get vaccinated?

- ○ Because of the coronavirus pandemic (give details below if you wish) (2) ________________________________________________
- ○ For another reason (give details below if you wish) (1) ________________________________________________

---

*Display This Question:*

*If Have you been vaccinated against whooping cough (also known as pertussis or Tdap) during your pre... = Yes*

Q18 Where did you receive your vaccine?

- ○ GP (1)
- ○ Antenatal care (hospital) (2)
- ○ Antenatal care (community setting) (3)
- ○ Pharmacy (4)
- ○ Other (5) ________________________________________________

---

*Display This Question:*

*If Do you plan to be vaccinated against whooping cough (also known as pertussis or Tdap) during your... = Yes*

*Or Do you plan to be vaccinated against whooping cough (also known as pertussis or Tdap) during your... = Unsure but leaning towards yes*

Q49 Where do you plan to receive your vaccine?

- ◯ GP (1)
- ◯ Antenatal care (hospital) (2)
- ◯ Antenatal care (community setting) (3)
- ◯ Pharmacy (4)
- ◯ Other (5) ________________________________________________

---

Q26 Have you had a baby before, in the last 8 years (between 2012-2020)?

- ◯ Yes (1)
- ◯ No (2)

---

*Display This Question:*

*If Have you had a baby before, in the last 8 years (between 2012-2020)? = Yes*

Q48 The next set of questions is about your **previous** most recent pregnancy:

---

*Display This Question:*

*If Have you had a baby before, in the last 8 years (between 2012-2020)? = Yes*

Q43 Were you vaccinated against whooping cough (also known as pertussis or Tdap) during your **previous** pregnancy?

- ◯ Yes (1)
- ◯ No (2)

---

*Display This Question:*

*If Were you vaccinated against whooping cough (also known as pertussis or Tdap) during your previous... = No*

Q38 During your previous pregnancy, which of the following best describes why you were not vaccinated against whooping cough?

- ◯ I didn't know about the vaccine (1)
- ◯ I didn't want to be vaccinated (2)
- ◯ I wanted to be vaccinated, but didn't (provide more details if you wish) (3) ________________________________________________
- ◯ Other (provide more details if you wish) (4) ________________________________________________

---

*Display This Question:*

*If During your previous pregnancy, which of the following best describes why you were not vaccinated... = I didn't know about the vaccine*

Q44 If you had known about the vaccine, do you think you would have got vaccinated?

- ◯ Yes, definitely (1)
- ◯ Unsure but leaning towards yes (2)
- ◯ Unsure but leaning towards no (3)
- ◯ No, definitely not (4)

---

*Display This Question:*

*If Were you vaccinated against whooping cough (also known as pertussis or Tdap) during your previous... = Yes*

Q52 Where did you receive your whooping cough vaccine in your **previous** pregnancy?

- ○ GP (1)
- ○ Antenatal care (hospital) (2)
- ○ Antenatal care (community setting) (3)
- ○ Pharmacy (4)
- ○ Other (5) ________________________________________________

**End of Block: Currently pregnant block (branch displayed if currently pregnant selected)**

---

**Start of Block: Current/most recent pregnancy COVID impact on access/knowledge**

Q22 Have you had any appointments changed due to the coronavirus (COVID-19) pandemic? Leave blank if no changes.

| | Postponed by me (1) | Postponed by GP/hospital (2) | Cancelled by me (3) | Cancelled by GP/hospital (4) | Changed to telephone/online appointment (5) |
|---|---|---|---|---|---|
| An appointment at your GP surgery about your pregnancy (1) | ☐ | ☐ | ☐ | ☐ | ☐ |
| An appointment at your GP surgery for another reason (2) | ☐ | ☐ | ☐ | ☐ | ☐ |
| An appointment at your GP surgery to receive a vaccine during pregnancy (3) | ☐ | ☐ | ☐ | ☐ | ☐ |
| An appointment at the hospital to receive a vaccine during pregnancy (6) | ☐ | ☐ | ☐ | ☐ | ☐ |
| Any other antenatal appointment at the hospital (4) | ☐ | ☐ | ☐ | ☐ | ☐ |

Q53 Feel free to add any additional comments here:

________________________________________________________________

________________________________________________________________

________________________________________________________________

________________________________________________________________

________________________________________________________________

**End of Block: Current/most recent pregnancy COVID impact on access/knowledge**

**Start of Block: Impact of COVID on maternity vaccines**

Q24 Please select your level of agreement with the following statements

| | Strongly agree (1) | Somewhat agree (2) | Neither agree nor disagree (3) | Somewhat disagree (4) | Strongly disagree (5) | N/A (6) |
|---|---|---|---|---|---|---|
| The COVID pandemic has restricted my **physical access** to vaccines during pregnancy (1) | o | o | o | o | o | o |
| The COVID pandemic has made me feel **less safe** about going to get my vaccines during pregnancy (2) | o | o | o | o | o | o |

Q25 During the COVID pandemic, where would you have liked to receive your vaccines during pregnancy?

| | Like a great deal (1) | Like somewhat (2) | Neither like nor dislike (3) | Dislike somewhat (4) | Dislike a great deal (5) |
|---|---|---|---|---|---|
| GP surgery (1) | ○ | ○ | ○ | ○ | ○ |
| Antenatal appointment (hospital) (2) | ○ | ○ | ○ | ○ | ○ |
| Antenatal appointment (community setting) (3) | ○ | ○ | ○ | ○ | ○ |
| Pharmacy (4) | ○ | ○ | ○ | ○ | ○ |
| A drive-through/walk through outside service (5) | ○ | ○ | ○ | ○ | ○ |
| Other (6) | ○ | ○ | ○ | ○ | ○ |

Q54 Feel free to add any additional comments here:

________________________________________________________________

________________________________________________________________

________________________________________________________________

________________________________________________________________

________________________________________________________________

**End of Block: Impact of COVID on maternity vaccines**

**Start of Block: Sources of information**

Q28 Have you obtained information from any of the below sources about vaccinations for pregnant people during the coronavirus (COVID-19) pandemic? Please tick all that apply

- ☐ A face-to-face conversation with my GP or practice nurse (1)
- ☐ A telephone/video call with my GP or practice nurse (2)
- ☐ A face to face conversation with a midwife or obstetrician (3)
- ☐ A telephone/video conversation with a midwife or obstetrician (4)
- ☐ A letter sent to my home (5)
- ☐ A leaflet (6)
- ☐ NHS website (10)
- ☐ Other website/s (7) ________________________________________________
- ☐ Social media (8) ________________________________________________
- ☐ An app (e.g. MatImms or BabyBuddy) (9) ________________________________________________
- ☐ Other (11) ________________________________________________

---

Q27 **Who** would you like to get information from about vaccinations in pregnancy during the coronavirus pandemic? Please rank them from the one you like the most (1) to the one you like the least (8)

______ GP (1)
______ Midwife or Nurse (2)
______ Other health care professional (3)
______ Pharmacists (4)
______ Scientists (6)
______ Other trained professional (e.g. 111 service) (7)
______ Friends or family (8)
______ Other (9)

Q29 How would you like to get information from about vaccinations in pregnancy during the coronavirus pandemic? Please rank them from the one you like the most (1) to the one you like the least (9)

______ A face to face conversation (1)

______ A telephone/video call (2)

______ A letter sent to my home (5)

______ A leaflet (6)

______ NHS website (7)

______ Other website/s (10)

______ Social media (8)

______ An app (e.g. MatImms or BabyBuddy) (9)

______ Other (11)

**End of Block: Sources of information**

---

**Start of Block: View on paeds vaccines and COVID vaccine**

Q30 Please select how much you agree or disagree with the following statements regarding your unborn child or new baby:

| | Strongly agree (1) | Agree a little (2) | Neither agree nor disagree (3) | Disagree a little (4) | Strongly disagree (5) |
|---|---|---|---|---|---|
| The coronavirus (COVID-19) pandemic will make it physically difficult to get my baby vaccinated (1) | ○ | ○ | ○ | ○ | ○ |
| During the coronavirus (COVID-19) pandemic, I feel it is **important** to get my baby vaccinated (2) | ○ | ○ | ○ | ○ | ○ |
| During the coronavirus (COVID-19) pandemic, I feel it is **safe** to go to get my baby vaccinated (3) | ○ | ○ | ○ | ○ | ○ |

Q36 Please select how much you agree or disagree with the following statements about a future vaccine to protect against COVID-19:

| | Yes, definitely (1) | Unsure but leaning towards yes (2) | Unsure but leaning towards no (3) | No, definitely not (4) |
|---|---|---|---|---|
| If a vaccine against coronavirus (COVID-19) becomes available, I would get vaccinated **whilst pregnant** (1) | ○ | ○ | ○ | ○ |
| If a vaccine against coronavirus (COVID-19) becomes available, I would get vaccinated **whilst not pregnant** (2) | ○ | ○ | ○ | ○ |
| If a vaccine against coronavirus (COVID-19) becomes available, I would vaccinate my **baby** (3) | ○ | ○ | ○ | ○ |

Q55 Feel free to add any additional comments here:

________________________________________________________________

________________________________________________________________

________________________________________________________________

________________________________________________________________

________________________________________________________________

End of Block: View on paeds vaccines and COVID vaccine

Start of Block: End of survey

Q58
Do you have anything further you would like us to know about your feelings or experience of vaccinated during the current coronavirus (COVID-19) pandemic? If yes, please use the text box below:

________________________________________________________________

________________________________________________________________

________________________________________________________________

________________________________________________________________

________________________________________________________________

Q62 Did you find the survey easy to navigate? (add comments if you wish)

- ○ Yes (1) ________________________________________________
- ○ No (3) ________________________________________________

Q61

**Follow up telephone interview**

If you would like to help us further and would be willing to participate in an approximately 20-30 min long conversation on similar topics as that covered in this survey please provide your email address or mobile phone details in the boxes provide below.

Please note that if you provide us with your contact details, then your survey responses will be no longer anonymous.

As a thank you for taking part in an interview, you will receive a £10 shopping voucher that can be used either in-store or online.

- ○ E-mail (1) ________________________________________________
- ○ Phone (2) ________________________________________________

---

Q60

**Thank you for completing this survey!**

For more information about this research please feel free to contact us at any time and we will be happy to answer your questions. You can reach us at:
Sara Barnett (research midwife)
Beth Holder (lead investigator)

---

Q61

**You can close this browser window, or click the arrow below to end the survey. Thank you again for your time.**

**End of Block: End of survey**

---

**Start of Block: Not currently pregnant block (branch displayed if not currently preg selected)**

Q61 In this most recent pregnancy, did you experience pregnancy loss? (miscarriage, termination or still birth)

○ No (1)

○ Yes (2)

*Display This Question:*

*If In this most recent pregnancy, did you experience pregnancy loss? (miscarriage, termination or st... = Yes*

Q62 We are very sorry for your loss. You may find that you are unable to answer some of the questions in this survey. You are free to skip any questions you cannot answer, or do not wish to answer.

You are also welcome to add any further comments/views in any of the text boxes.

We thank you for taking the time to take part in this survey.

*Display This Question:*

*If In this most recent pregnancy, did you experience pregnancy loss? (miscarriage, termination or st... = No*

Q41 When did you give birth?

▼ March 2020 (1) ... December 2020 (10)

Q54 Whooping cough vaccine (also known as pertussis or Tdap) is recommended to all pregnant people in England. Did you know this?

○ Yes (1)

○ No (2)

*Display This Question:*

*If Whooping cough vaccine (also known as pertussis or Tdap) is recommended to all pregnant people in... = No*

Q43 If you had known this vaccine was recommended in pregnancy, would you have been vaccinated?

- ◯ Yes (1)
- ◯ Unsure but leaning towards yes (2)
- ◯ Unsure but leaning towards no (3)
- ◯ No (4)

---

*Display This Question:*

*If Whooping cough vaccine (also known as pertussis or Tdap) is recommended to all pregnant people in... = Yes*

Q55 Were you vaccinated against whooping cough (also known as pertussis or Tdap) during your pregnancy?

- ◯ Yes (1)
- ◯ No (2)

---

*Display This Question:*

*If Were you vaccinated against whooping cough (also known as pertussis or Tdap) during your pregnancy? = No*

Q60 Which of the following best describes why you were not vaccinated during your pregnancy?

- ◯ I didn't want to be vaccinated (2)
- ◯ I didn't know about the vaccine (1)
- ◯ I wanted to be vaccinated but didn't because of the COVID-19 pandemic (give details below if you wish) (5) ________________________________________________
- ◯ I wanted to be vaccinated but didn't for a different reason (give details below if you wish) (4) ________________________________________________
- ◯ Other (3) ________________________________________________

---

*Display This Question:*

*If Which of the following best describes why you were not vaccinated during your pregnancy? = Other*

Q61 If you selected Other, please provide more information if you wish:

________________________________________________________________

---

*Display This Question:*

*If Which of the following best describes why you were not vaccinated during your pregnancy? = I didn't know about the vaccine*

Q45 If you had known this vaccine was recommended in pregnancy, would you have been vaccinated?

- ◯ Yes (1)
- ◯ Unsure but leaning towards yes (2)
- ◯ Unsure but leaning towards no (3)
- ◯ No (4)

---

*Display This Question:*

*If Were you vaccinated against whooping cough (also known as pertussis or Tdap) during your pregnancy? = Yes*

Q56 Where did you receive your vaccine?

- ◯ GP (1)
- ◯ Antenatal care (hospital) (2)
- ◯ Antenatal care (community setting) (3)
- ◯ Pharmacy (4)
- ◯ Other (5) ________________________________________________

---

Q58 Have you had another baby before, in the last 8 years (between 2012-2020)?

- ◯ Yes (1)
- ◯ No (2)

---

*Display This Question:*

*If Have you had another baby before, in the last 8 years (between 2012-2020)? = Yes*

Q47 The next set of questions is about your **previous** pregnancy, before your most recent one

---

*Display This Question:*

*If Have you had another baby before, in the last 8 years (between 2012-2020)? = Yes*

Q66 Were you vaccinated against whooping cough (also known as pertussis or Tdap) during your **previous** pregnancy?

- ◯ Yes (1)
- ◯ No (2)

---

*Display This Question:*

*If Were you vaccinated against whooping cough (also known as pertussis or Tdap) during your previous... = No*

Q67 During your previous pregnancy, which of the following best describes why you were not vaccinated against whooping cough?

- ◯ I didn't know about the vaccine (1)
- ◯ I didn't want to be vaccinated (2)
- ◯ Other (3)

---

*Display This Question:*

*If During your previous pregnancy, which of the following best describes why you were not vaccinated... = Other*

Q68 If you selected Other, please provide more information if you wish:

________________________________________________________________

---

*Display This Question:*

*If During your previous pregnancy, which of the following best describes why you were not vaccinated... = I didn't know about the vaccine*

Q46 If you had known this vaccine was recommended, would you have been vaccinated in your **previous** pregnancy?

- ◯ Yes (1)
- ◯ Unsure but leaning towards yes (2)
- ◯ Unsure but leaning towards no (3)
- ◯ No (4)

---

*Display This Question:*

*If Were you vaccinated against whooping cough (also known as pertussis or Tdap) during your previous... = Yes*

Q69 Where did you receive your whooping cough vaccine in your **previous** pregnancy?

- ◯ GP (1)
- ◯ Antenatal care (hospital) (2)
- ◯ Antenatal care (community setting) (3)
- ◯ Pharmacy (4)
- ◯ Other (5) ________________________________________________

**End of Block: Not currently pregnant block (branch displayed if not currently preg selected)**
