## Supplementary B for "Women’s views and experiences of accessing vaccination in pregnancy during the COVID-19 pandemic: A multi-methods study in the United Kingdom"

Imperial College London

#### Participant Information Sheet

### Women's views and experience of maternal vaccination during the coronavirus (COVID-19) pandemic

Study Group
Principal Investigator: Dr Beth Holder

Co-investigators:
*Imperial College London:*
Dr Helen Skirrow
Ms Sara Barnett

*London School of Hygiene & Tropical Medicine (LSHTM)*
Prof. Beate Kampmann
Dr Sadie Bell
Assoc. Prof. Sandra Mounier-Jack

**INTRODUCTION**

Thank you for completing our online survey on the impact of the COVID-19 pandemic on pregnant women's views and access to maternal vaccines. We would like to invite you to take part in a follow up study which involves a short telephone interview. This leaflet provides information to help you decide whether or not you would like to take part in the study – taking part is completely voluntary.

Please take your time to read the following information carefully and to talk to others about the study, if you wish. Ask us (the study researchers) if there is anything that is not clear or if you would like more information.

Please ask us if there is anything that is not clear or if you would like more information.
Take time to decide whether or not you wish to take part.

Thank you for reading this.

- **What is the purpose of the study?**

The purpose of this study is to learn about women's views on maternal vaccination and their experiences of using GP and hospital services for vaccination during the coronavirus (COVID-19) pandemic. This will help inform the way that maternal vaccinations are communicated and delivered during pandemics.

- **Why have I been invited?**

We would like to interview women, aged 16 years and over, who have been pregnant at any time during the UK coronavirus pandemic (since 23rd March). You have been invited to take part as you have completed our online survey and have expressed an interest in taking part in a follow-up interview. We will interview 20-40 women.

- **Do I have to take part?**

It is up to you to decide whether or not to take part. If you do decide to take part you will be given this information sheet to keep and our research midwife will complete a consent form with you over the telephone. You will be provided with a copy of this consent form. If you do decide to take part and later change your mind, you can withdraw up to two weeks after your interview. You do not have to give a reason. We will delete your interview. After this time, your interview will have been transcribed and anonymised, and it will not be possible to delete it.

- **What will happen to me if I take part?**

You indicated on the survey that you would potentially like to take part in a follow up interview. This would involve one of the study researchers contacting you via telephone to take part in an audio-recorded interview lasting around 20-30 minutes. If you decide to take part, we will ask you to provide your verbal or written consent for this study as a record of your decision to take part in the study. A copy of the written information and signed Informed Consent form will be given to you by e-mail/post to keep.

In the interview we would like to talk to you about whether your GP practice or the hospital providing your antenatal care have contacted you about vaccinations during pregnancy. We would like to ask you about how you feel about attending your GP practice or antenatal appointments to get vaccinated during the coronavirus pandemic. If you did get vaccinated in your pregnancy, we would like to hear about your experience of this. If you did not get vaccinated in your pregnancy, we would also like to hear your views on this. Finally, we would also like to ask you about your views on getting your new baby vaccinated, and about potential future coronavirus vaccines.

The audio-recordings from the interview will be transcribed into text, and anonymised so that the people taking part in the interview cannot be identified. The audio-recording will then be deleted. We will store the interview data securely in line with Research Ethics Committee guidelines and only the study researchers will have access to this. Data will be stored in password-protected files in Imperial College's secure OneDrive account. We may use quotes from the interviews in reports and academic publications but these will be anonymous (no names recorded).

At the end of the project, the study data will be archived at Imperial College London. The data will be made available to other UK researchers for research and to improve healthcare, if you consent to this. Your personal information will not be included and there is no way that you can be identified.

All information collected about you will be kept strictly confidential. The decision about whether or not you take part is up to you. You are under no obligation to participate.

- **What do I have to do?**

You will need to provide the research midwife with suitable times for her to contact you to carry out the interview. The interview should take around 20-30 minutes.

- **What are the possible disadvantages and risks of taking part?**

There are no predicted disadvantages nor risks of taking part. The interview is intended to be an opportunity for you to express your views in a non-judgemental space. You are free to skip any question that you do not want to answer and can withdraw from the interview at any time. If you feel uncomfortable at any time, please let us know and we can stop the interview.

- **What are the possible benefits of taking part?**

Taking part in the study is unlikely to benefit you directly, however the information you share with us will help inform the way that maternal vaccinations are communicated and delivered during pandemics.

To thank you for taking the time to participate, we will offer you a £10 gift voucher.

- **What if something goes wrong?**

If you are harmed by taking part in this research project, there are no special compensation arrangements. If you are harmed due to someone's negligence, then you may have grounds for a legal action. Regardless of this, if you wish to complain, or have any concerns about any aspect of the way you have been treated during the course of this study then you should immediately inform the Investigator Beth Holder. If you are still not satisfied with the response, you may contact the Imperial Joint Research Compliance Office.

- **What will happen to the results of the research study?**

When the study is completed, we will produce a report of what participants have told us. If you would like, we will send you a summary of this report. Study findings will be presented at relevant meetings and conferences and will be published within peer-reviewed academic journals.

All information collected about you will be kept strictly confidential. Your name and any other information that could lead to your identity being revealed will not appear in any reports or publications.

- **Who is organising and funding the research?**

This research is organised by Imperial College London, Department of Metabolism, Digestion and Reproduction. It is funded by the Imperial College COVID Research Fund.

- **Who has reviewed the study?**

This study was reviewed by the Imperial College COVID19 Research Fund and the Imperial College Academic Health Sciences Centre COVID19 Research Committee. It was given approval by the Head of Department and the Imperial College Joint Research Compliance Office (JRCO).

**Contact for Further Information**

Study phone number: 07999 042842

You are encouraged to present any concerns or complaints to either Sara Barnett (see contact details above) or to: Beth Holder, Lecturer in Maternal and Fetal Health, Imperial College, IRDB, Hammersmith Campus, W12 0HS,.

Thank you for taking part in this study.

Imperial College London

**TRANSPARENCY NOTICE**

**HOW WILL WE USE INFORMATION ABOUT YOU?**

**Research Study Title: Women's views and experience of maternal vaccination during the coronavirus (COVID-19) pandemic**

Imperial College London is the sponsor for this study and will act as the data controller for this study. This means that we are responsible for looking after your information and using it properly. Imperial College London will keep your personal data for:

- 10 years after the study has finished in relation to data subject consent forms.
- 10 years after the study has completed in relation to primary research data.

We will need to use information from you for this research project.

This information will include your name and contact details. People will use this information to do the research or to check your records to make sure that the research is being done properly.

People who do not need to know who you are will not be able to see your name or contact details. Your data will have a code number instead.

We will keep all information about you safe and secure.

Once we have finished the study, we will keep some of the data so we can check the results. We will write our reports in a way that no-one can work out that you took part in the study.

**LEGAL BASIS**

As a university we use personally-identifiable information to conduct research to improve health, care and services. As a publicly-funded organisation, we have to ensure that it is in the public interest when we use personally-identifiable information from people who have agreed to take part in research. This means that when you agree to take part in a research study, we will use your data in the ways needed to conduct and analyse the research study. Health and care research should serve the public interest, which means that we have to demonstrate that our research serves the interests of society as a whole. We do this by following the UK Policy Framework for Health and Social Care Research

**INTERNATIONAL TRANSFERS**

There may be a requirement to transfer information to countries outside the European Economic Area (for example, to a research partner). Where this information contains your personal data, Imperial College London will ensure that it is transferred in accordance with data protection legislation. If the data is transferred to a country which is not subject to a European Commission (**EC**) adequacy decision in respect of its data protection standards, Imperial College London will enter into a data sharing agreement with the recipient organisation that incorporates EC approved standard contractual clauses that safeguard how your personal data is processed.

**SHARING YOUR INFORMATION WITH OTHERS**

For the purposes referred to in this privacy notice and relying on the bases for processing as set out above, we will share your personal data with certain third parties.

- Other College employees, agents, contractors and service providers (for example, suppliers of printing and mailing services, email communication services or web services, or suppliers who help us carry out any of the activities described above). Our third party service providers are required to enter into data processing agreements with us. We only permit them to process your personal data for specified purposes and in accordance with our policies.
- the following Research Collaborators / Partners in the study;
  - London School of Hygiene and Tropical Medicine (LSHTM) – the transcribed interview will be shared with our co-investigators at LSHTM. These co-investigators are experts in social science and public health and will contribute to the analysis of the results.

**WHAT ARE YOUR CHOICES ABOUT HOW YOUR INFORMATION IS USED?**

You can stop being part of the study at any time, without giving a reason. Within two weeks of the interview, we can delete the interview. After two weeks, your interview will have been transcribed and anonymised, and therefore we will not be able to delete it.

- We need to manage your records in specific ways for the research to be reliable. This means that we won't be able to let you see or change the data we hold about you.
- If you agree to take part in this study, you will have the option to take part in future research using your data saved from this study.

**WHERE CAN YOU FIND OUT MORE ABOUT HOW YOUR INFORMATION IS USED**

You can find out more about how we use your information

- by asking one of the research team
- www.theholderlab.com/info

**COMPLAINT**

If you wish to raise a complaint on how we have handled your personal data, please contact Imperial College London's Data Protection Officer via email at, via telephone on 020 7594 3502 and/or via post at Imperial College London, Data Protection Officer, Faculty Building Level 4, London SW7 2AZ.

If you are not satisfied with our response or believe we are processing your personal data in a way that is not lawful you can complain to the Information Commissioner's Office (ICO). The ICO does recommend that you seek to resolve matters with the data controller (us) first before involving the regulator.

Imperial College London

### Consent Form for Participants Able to Give Consent

**Full Title of Project: Women's views and experience of maternal vaccination during the coronavirus (COVID-19) pandemic**

Name of Principal Investigator: Dr Beth Holder

Consent can either be completed by e-mail or over the telephone. If possible, please complete this and return by e-mail prior to your interview. Initial each of the boxes to indicate whether you agree with the statements, and then type your name and the date at the bottom. **Please also confirm in the e-mail that you consent to all the points.** Alternatively, the research midwife can complete it with you over the phone before the interview starts.

**Please put your initials in each of these boxes**

| |
|---|
| 1. I confirm that I have read and understand the participant information sheet version 1.1. dated 22/07/2020 for the above study and have had the opportunity to ask questions which have been answered fully. |
| 2. I understand that my participation is voluntary, and I am free to withdraw at any time, without giving any reason and without my legal rights being affected. |
| 3. I give permission for Imperial College London to access my records that are relevant to this research. |
| 4. I understand that I am free to answer any questions that I do not wish to answer. |
| 5. I understand that my interview will be audiorecorded. |
| 6. I consent to take part in the above study. |
| 7. I give/do not give (delete as applicable) consent for information collected about me to be used to support other research in the future, including those outside of the European Economic Area (EEA). |
| 8. I give/do not give (delete as applicable) consent to being contacted to potentially taking part in other research studies (optional) |

| | | |
|---|---|---|
| ______________________ | ______________________ | ______________________ |
| Name of Participant | Signature (type your name here) | Date |
| ______________________ | ______________________ | ______________________ |
| Name of Person taking consent (if different from Principal Investigator) | Signature | Date |
| ______________________ | ______________________ | ______________________ |
| Principal Investigator | Signature | Date |
