## Supplementary C for "Women’s views and experiences of accessing vaccination in pregnancy during the COVID-19 pandemic: A multi-methods study in the United Kingdom"

Imperial College London

**Covid Survey 2020: Topic guide for Interviews**

- *Introduction and recap the purpose of the study.*
- *Acknowledge that the interview will be anonymous, and no identifiable information will be shared with those outside of the study.*
- *Responses will be shared anonymously and may be quoted in publications.*

*(Data to completed as far as possible from survey info)*

| | | |
|---|---|---|
| Name / Participant number | | |
| Date | | |
| Time | | |
| Email | | |
| Telephone | | |
| Age | | |
| Geographic Area | | |
| Employment status | | |
| Ethnicity | | |
| Date of Birth of new Baby | | |
| Number of children | | |
| Ages of children | | |
| Immunisation status in this pregnancy | Pertussis | Flu |
| Immunisation status in previous pregnancies | Pertussis | Flu |
| If still pregnant - how many weeks? | | |
| Any health complications during pregnancy | | |

1. **Can you tell me about your experience of being pregnant during the corona virus period?**

Prompts

- How different from previous pregnancies
- Different antenatal care?
- Partner attendance at appts?

2. **Thinking about your pregnancy can you please tell me about your experience of vaccination?**

Prompts

- Did you know you should be vaccinated during pregnancy?
  - Whooping cough / Flu
- Do you know why these vaccines are recommended?
- Please tell me how you learned that you should receive vaccinations during your
- pregnancy?
- Who discussed vaccination with you? And how? i.e. letter, text msg, verbal
  - When did this occur?
  - Given enough info to make informed decision?
  - Given opportunity to ask any questions?
  - Were you able to get vaccinated?
- How easy was it to access the centre where the vaccines were administrated?
  - making an appointment,
  - experience of the appointment
- Why did you decide to/not to vaccinate?
  - (was the decision an active and considered choice or simply following advice?)
- What do you think are the most important influences to your decision to vaccinate?
  - Who? Role of family/friends
- Would you recommend vaccination in pregnancy to friends/relatives?

**3. Tell me about your new baby …..**

Prompts

- Have they started/completed their vaccination schedule?
- And other children?
- If <8 weeks, do they plan to fully vaccinate their baby?

If no, explore in what way they plan to modify and reasons.

**4. There has been much discussion of a vaccine against Covid**

- What are your feelings about receiving the vaccine?

Prompts

- Would you have the vaccine?
    - When not pregnant/when pregnant
- Would you give it to your baby/child?
- What are your concerns ….

*Thank you*

*I have no further questions, is there anything we have not discussed that you would like to tell me more about.*

*Is there anything you would like to ask me?*
