## Supplementary Figures and Tables for "Women’s views and experiences of accessing vaccination in pregnancy during the COVID-19 pandemic: A multi-methods study in the United Kingdom"

### Supplementary Figure 1: Demographics Survey Respondents

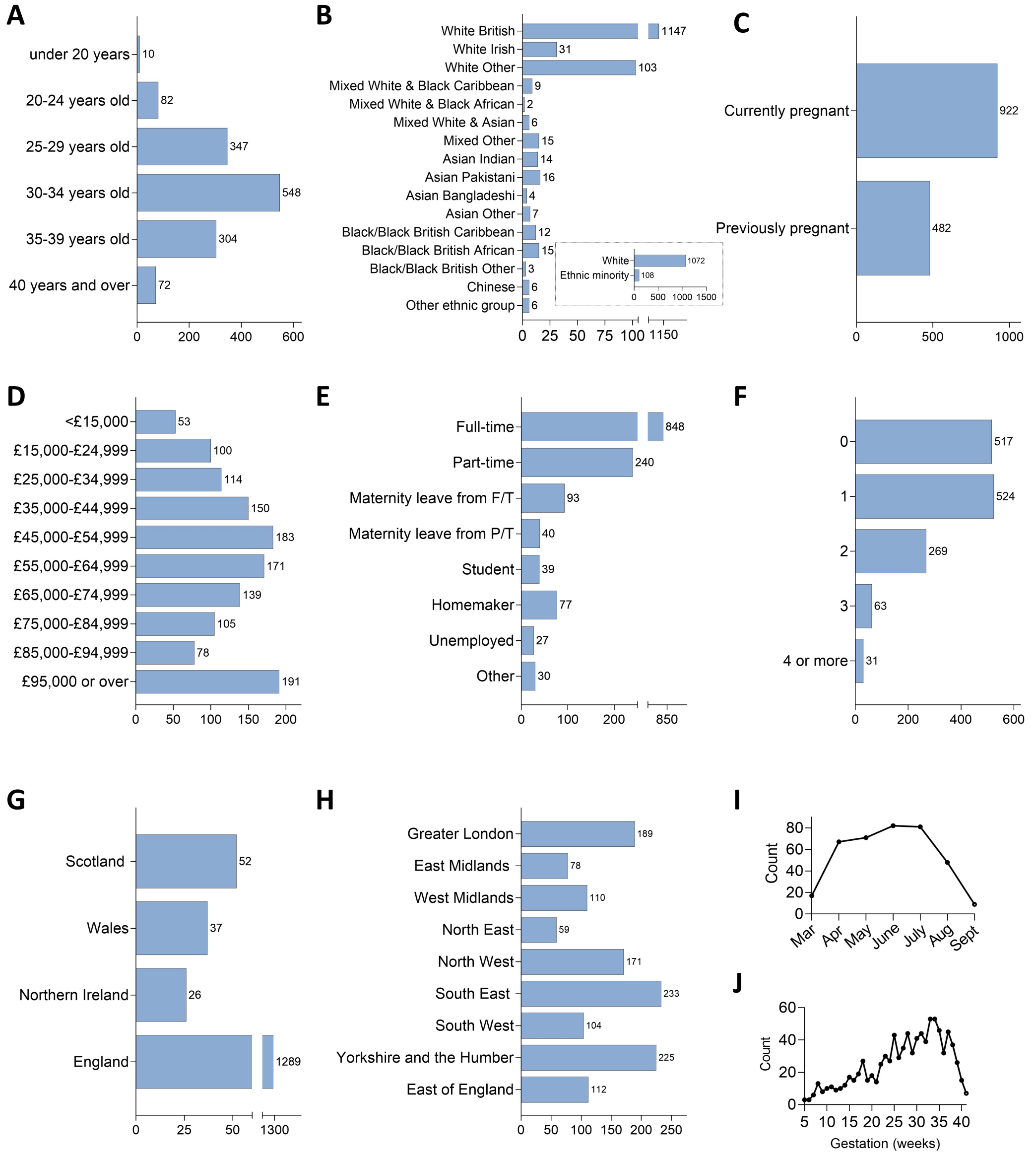

**Supplementary Figure 1: Demographics of Survey Respondents.** Self-reported demographics of the survey respondents at the time of survey completion. These questions were **optional.** A) Age (years); B) Ethnicity; C) Pregnancy status; D) Annual household income (£); E) Employment status; F) Number of children  G) Country of residence; H) Region of residence for English residents; I) Month of delivery for new mothers; J)Gestational age of pregnancy (weeks) for those currently pregnant;.

### *Supplementary Figures and Tables*

#### Supplementary Table 1: Interviewee Characteristics

| ID | Ethnicity | Parity | Pertussis vaccine in pregnancy |
|---|---|---|---|
| 1 | Chinese | 3 | Yes |
| 2 | White Asian | 1 | Yes |
| 3 | British Pakistani | 1 | No |
| 4 | British Arabic | 1 | Yes |
| 5 | Black-African | 1 | Yes |
| 6 | White British | 2 | Yes |
| 7 | White British | 4 | Yes |
| 8 | White British | 2 | Yes |
| 9 | White British | 2 | Yes |
| 10 | White British | 1 | Yes |

#### Supplementary Table 2: Uptake of pertussis vaccine in pregnancy

| **Have you been vaccinated against whooping cough (also known as pertussis or Tdap) during your pregnancy?** | | |
|---|---|---|
| | **VACCINATED**<br><br>**n (%)** | **UNVACCINATED**<br><br>**n (%)** |
| **Women pregnant at the time of the survey (all)** | **624 (72.1)** | **241 (27.9)** |
| *First trimester* | *2 (0.3)* | *57 (23.7)* |
| *Second trimester* | *158 (25.3)* | *142 (58.9)* |
| *Third trimester* | *464 (74.4)* | *42 (17.4)* |
| **Women who had delivered at the time of the survey** | **408 (89.1)** | **50 (10.9)** |

#### Supplementary Table 3: Intentions to be vaccinated among pregnant unvaccinated survey respondents

| **Do you plan to be vaccinated against whooping cough (also known as pertussis or Tdap) during your pregnancy?** | | |
|---|---|---|
| | **Y**<br>**n (%)** | **N**<br>**n (%)** |
| **Women pregnant at the time of the survey unvaccinated against pertussis** | **204 (84.6)** | **37 (15.4)** |

Women who were currently pregnant at the time of the survey and answered that they were unvaccinated were asked *'Do you plan to be vaccinated against whooping cough (also known as pertussis or Tdap)?'. Possible answers were: 'Yes', 'Unsure but leaning towards yes', 'Unsure but leaning towards no' and 'No'. 'Yes' (n=181) and 'Unsure but leaning towards yes' (n=23) combined into* ***Y*** *and 'Unsure but leaning towards no' (n=7) and 'No' (n=30) combined into* ***N****.*

#### Supplementary Table 4: Intentions to be vaccinated among survey respondents unaware that pertussis vaccine in pregnancy.

| **Do you plan to be vaccinated against whooping cough (also known as pertussis or Tdap) during your pregnancy?** | | |
|---|---|---|
| | **Y**<br>**n (%)** | **N**<br>**n (%)** |
| **Women pregnant at the time of the survey but unaware of pertussis vaccine recommended** | **35 (76.0)** | **11 (24.0)** |

### *Supplementary Figures and Tables*

Women who were currently pregnant at the time of the survey and answered that they were not aware that pertussis vaccine was recommended in pregnancy were asked: *'Now you know Tdap is recommended do you think you will get vaccinated?'*

### Supplementary Table 5: Reasons for being unvaccinated in pregnancy

| | **Pregnant at time of survey n (%)** | **Delivered at the time of survey n (%)** |
|---|---|---|
| *'I did not want to be vaccinated'* | - | 1 (2.1) |
| *'I didn't know about the vaccine'* | - | 20 (42.6) |
| *'I wanted to be vaccinated but didn't because of the COVID-19 pandemic'* | 22 (25.3) | 10 (21.3) |
| *'I wanted to be vaccinated but didn't for a different reason'* | - | 5 (10.6) |
| *'Other'* | 65 (74.7) | 11 (23.4) |

Women who were pregnant at the time of the survey and unvaccinated were asked '*Which of the following best describes why you are unsure or not planning to be vaccinated'*
Women who had already delivered at the time of the survey but were unvaccinated in pregnancy were asked *'Which of the following best describes why you were not vaccinated during your pregnancy?'.*

### Supplementary Table 6: Location where pertussis vaccine in pregnancy delivered

| **Where did you receive your vaccine?** | |
|---|---|
| **Location** | **n (%)** |
| General Practice | 637, (61.8%) |
| Antenatal care (Hospital) | 316, (30.7%) |
| Antenatal (Community) | 55, (5.3%) |
| Pharmacy | 2, (0.2%) |
| Other | 20, (1.9%) |

Women were asked *'Where did you receive your vaccine*

### Supplementary Table 7: Location where women planning to be vaccinated

| **Where do you plan to have your vaccine?** | |
|---|---|
| **Location** | **n (%)** |
| General Practice | 114, (55.9%) |
| Antenatal care (Hospital) | 59, (28.9%) |
| Antenatal (Community) | 21, (10.3%) |
| Pharmacy | 1, (0.50%) |
| Other | 9, (4.4%) |

**Women who were unvaccinated and pregnant were asked 'Where do you plan to have your vaccine?'.**

### Supplementary Figure 2: Healthcare Setting for delivery of vaccines to pregnant women before and during the pandemic, and where women prefer to be vaccinated

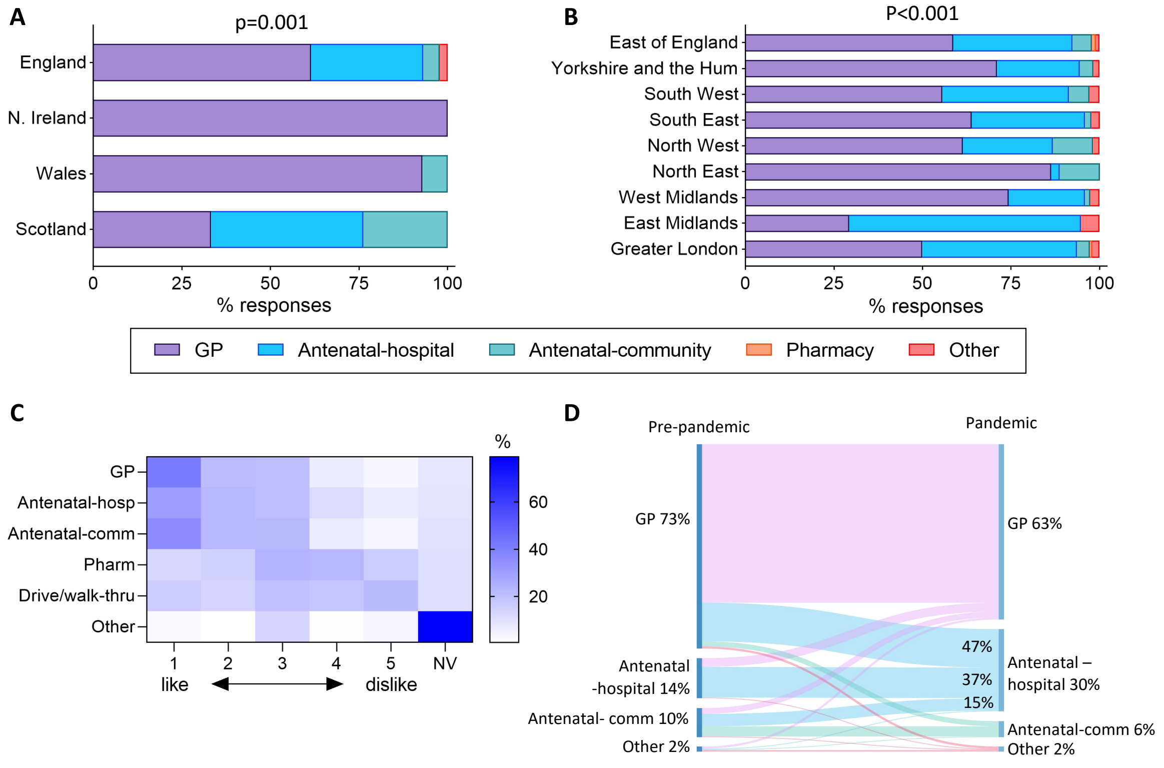

**Supplementary Figure 2. Healthcare setting for delivery of vaccines to pregnant women before and during the pandemic, and where women prefer to be vaccinated.** A) Location of pertussis vaccination delivery during pregnancy, separated by UK country. Analysed by Chi-Square Test. B) Location of pertussis vaccination delivery during pregnancy across English geographical regions. Analysed by Chi-Square Test. C) Preferred location for pregnancy vaccination during the COVID-19 pandemic. Women were asked 'During the COVID pandemic, where would you have liked to receive your vaccines during pregnancy'. Responses were scored on a Likert scale (Like a great deal, Like somewhat, Neither like or dislike, Dislike somewhat and Dislike a great deal). D) Sankey plot of multiparous women showing where they were vaccinated during the pandemic compared to their most recent previous pregnancy, prior to the pandemic. None of the women who had a prior pregnancy reported receiving their vaccine in a pharmacy (before nor during the pandemic). The % values represent the proportion of women who received their vaccine in the antenatal care hospital setting who had a previous vaccine given in the same (37%) or a different setting. Diagram created using SankeyMATIC. GP; general practitioners; Antenatal-hosp; antenatal care in hospital setting, Antenatal-comm; antenatal care in community setting, Pharm; pharmacy.

Supplementary Figure 3: Travel to antenatal appointments before and after the national lockdown from 23rd March 2020 in England.

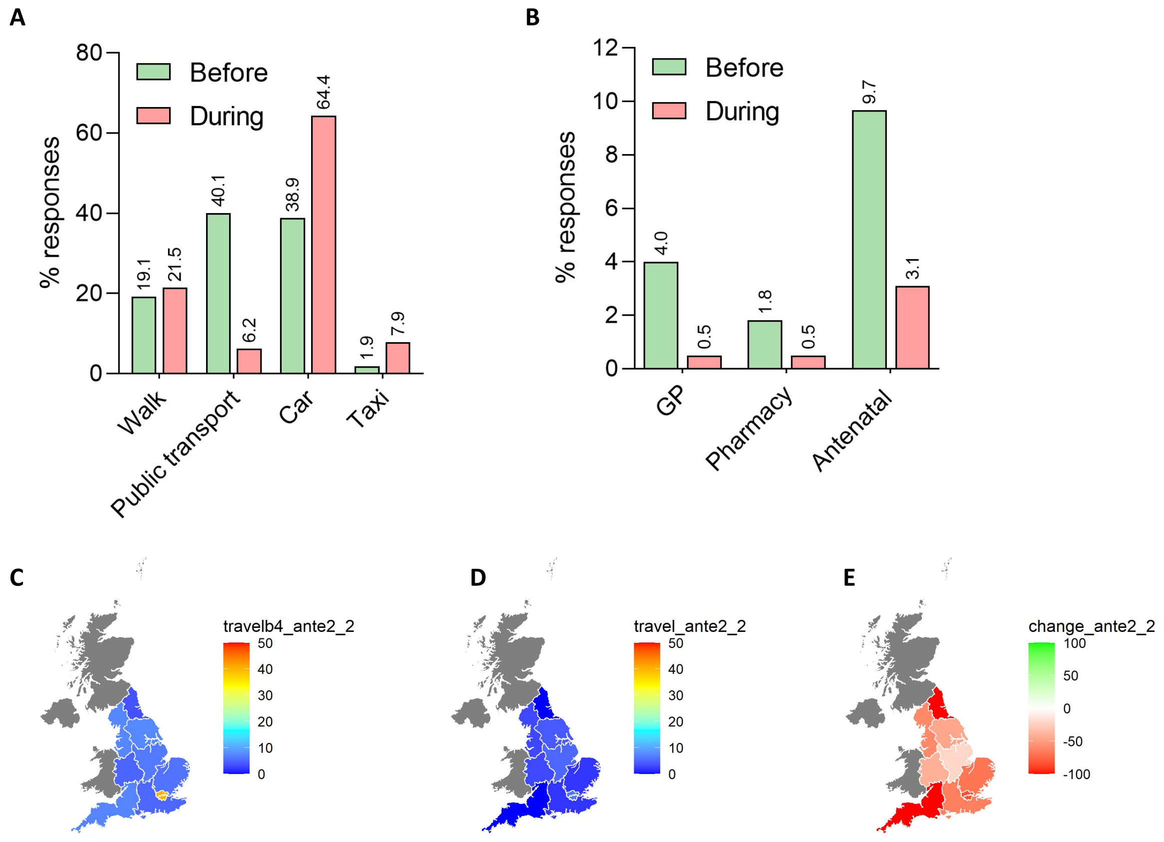

**Supplementary Figure 3:**
**Travel to antenatal appointments before and after the national lockdown from 23rd March 2020 in England.**

Women were asked 'Before the lockdown began (23rd March 2020), how would you usually get to the following: face-to-face appointments at their GP, pharmacy and antenatal hospital settings?' And then 'After the lockdown began (23rd March 2020), how have you usually got to the following: face-to-face appointments at their GP, pharmacy and antenatal hospital settings?'

Responses were walking, public transport, care or taxi.

A: Travel to antenatal appointments before and after the COVID-19 pandemic national lockdown response.
B: Travel to antenatal appointments by public transport at different settings before and after the COVID-19 pandemic national lockdown response.
C: Travel to antenatal appointments by public transport in England before the COVID-19 pandemic national lockdown response.
D: Travel to antenatal appointments by public transport in England after the COVID-19 pandemic national lockdown response.
E: % change in travel to antenatal appointments by public transport in England before and after the COVID-19 pandemic national lockdown response.
